# Burden and Genomic Landscape of Antimicrobial Resistance in Non-Bloodstream Infections Among Patients with Cirrhosis: A Systematic Review and Meta-Analysis

**DOI:** 10.64898/2025.12.12.25342176

**Authors:** Cansu Ozdemir, Tuhin Paul, Kasope Egoh, Joseph J. Wanford, Debbie L. Shawcross, Ellis K. Paintsil

## Abstract

**Background:** Patients with cirrhosis are highly susceptible to infections due to immune dysfunction and gut barrier impairment. Non-bloodstream infections frequently trigger decompensation and mortality, yet the global distribution of antimicrobial resistance (AMR) in these infections is poorly characterized.

**Methods:** We performed a systematic review and meta-analysis of studies reporting AMR in non-bloodstream bacterial infections among patients with cirrhosis. PubMed, Embase, and Web of Science were searched up to 15 September 2025. Pooled prevalence estimates of multidrug-resistant (MDR) and key resistant pathogens were calculated using random-effects models. Subgroup analyses were performed by country income, continent, and bacterial species.

**Results:** Thirty-one studies including 3,162 infections were analysed. Spontaneous bacterial peritonitis predominated (79%), followed by colonisation (12%) and urinary tract infections (7%). Gram-negative bacteria accounted for 60% of infections (Escherichia coli 29%, Klebsiella pneumoniae 11%), while Gram-positive pathogens represented 39% (Enterococcus spp. 14%, Staphylococcus aureus 6%). Overall pooled MDR prevalence was 29%, with higher burdens in lower–middle-income countries (MDR 47% vs. 22–41%; ESBL 24% vs. 10%; VRE 21% vs. 3%; CRE 32% vs. 1%). Genotypic data identified 436 resistance genes with marked continental differences.

**Conclusion:** Cirrhosis-associated non-bloodstream infections are dominated by Gram-negative bacteria and show high MDR, particularly in lower–middle-income countries. These findings highlight the need for integrated phenotypic and genomic surveillance of resistance patterns in these settings to guide empiric therapy.

## Background

Patients with cirrhosis are profoundly vulnerable to bacterial infections due to cirrhosis-associated immune dysfunction, impaired intestinal barrier integrity, and marked alterations in the gut microbiota (Liu et al., 2025; Noor & Manoria, 2017; Suárez et al., 2025; Tsiaoussis et al., 2015). These pathophysiological disturbances facilitate bacterial overgrowth and translocation, predisposing individuals to non-bloodstream infections—such as spontaneous bacterial peritonitis (SBP), urinary and respiratory infections—which are major triggers of decompensation, acute-on-chronic liver failure, and short-term mortality (Marciano et al., 2019; Piano & Angeli, 2021). Recurrent hospitalisation, invasive procedures, and frequent antibiotic exposure further compound this susceptibility, positioning infections as a leading driver of morbidity in advanced liver disease (Suárez et al., 2025).

At the same time, antimicrobial resistance (AMR) is escalating globally (Paintsil et al., 2025), undermining the effectiveness of standard empirical regimens used in cirrhosis. Multidrug-resistant gram-negative organisms, particularly *Escherichia coli* and *Klebsiella pneumoniae*, alongside resistant gram-positive pathogens, now frequently complicate clinical management (Macesic et al., 2025). Mechanistic evidence suggests that cirrhosis may foster both selection and propagation of multidrug-resistant organisms and antimicrobial resistance genes through gut dysbiosis, increased intestinal permeability, and repeated antibiotic exposure (Liu et al., 2025). This convergence of biological susceptibility and expanding global AMR highlights an urgent need for integrated phenotypic and genomic surveillance in this high-risk population.

Despite these intersecting risks, current understanding of pathogen distributions and resistance patterns in cirrhosis remains fragmented (Iskandar et al., 2021; Liakina, 2023). Existing studies predominantly focus on bloodstream infections, single organisms, or isolated geographic regions, with limited integration of phenotypic susceptibility profiles and genotypic determinants (Paintsil et al., 2025; Patel & Williams, 2019). Critically, non-bloodstream infections—by far the most frequent infections in cirrhosis—have not been comprehensively characterised across global populations, and data from regions with rising cirrhosis burden and limited diagnostic capacity remain scarce (Cao et al., 2024; Piano et al., 2019). To address these gaps, we conducted a systematic review and meta-analysis integrating bacterial pathogens, resistance phenotypes, and reported antimicrobial resistance genes across all major non-bloodstream infections in cirrhosis, providing a global framework to guide empiric therapy, antimicrobial stewardship, and international AMR preparedness.

## Methodology

### Study design and protocol registration

This systematic review and meta-analysis was conducted following the PRISMA-P 2020 guidelines for protocol preparation (Page, McKenzie, et al., 2021) and reported in accordance with the PRISMA 2020 statement for systematic reviews (Page, Moher, et al., 2021). The study protocol was prospectively registered in PROSPERO (CRD420251076038).

### Literature search

A comprehensive literature search was conducted on 15 September 2025 across MEDLINE (via PubMed), Embase (via Ovid), Scopus, and Web of Science, from inception to the search date. The search combined controlled vocabulary (e.g., MeSH/Emtree terms) and free-text keywords related to cirrhosis, antimicrobial resistance and resistance genes (ARGs), and non-bloodstream infections, including SBP, urinary tract infection (UTI), pneumonia, and soft tissue infections. Full search strategies for each database are provided in the Supplementary File 1. Titles and abstracts were independently screened by two reviewers (EK and TP) using Rayyan software, followed by full-text assessment of potentially eligible studies. Discrepancies were resolved through discussion or, if required, consultation with a third reviewer (CO).

### Inclusion Criteria

We included studies of adults (≥18 years) with a confirmed diagnosis of liver cirrhosis in whom bacterial pathogens were isolated and evaluated for antimicrobial resistance. Eligible studies reported either genotypic resistance, detecting antimicrobial resistance genes (ARGs) via molecular methods such as polymerase chain reaction (PCR) or whole-genome sequencing (WGS), or phenotypic resistance, including multidrug-resistant organisms (MDROs) such as *methicillin-resistant Staphylococcus aureus* (MRSA), *extended-spectrum β-lactamase*-producing *Escherichia coli* (ESBL), carbapenem-resistant Enterobacterales (CRE), vancomycin-resistant *Enterococcus* (VRE), and fluoroquinolone-resistant *Pseudomonas* species. Studies were required to focus on non-bloodstream infections, including spontaneous bacterial peritonitis, urinary tract infections, pneumonia, wound infections, or soft tissue infections. Studies reporting both bloodstream and non-bloodstream infections were eligible only if outcomes were clearly stratified by infection type. All observational studies (retrospective and prospective cohorts, case-control, cross-sectional) were considered, with no restrictions on geographic region, country income level, or publication year.

### Exclusion Criteria

We excluded studies that focused exclusively on bloodstream infections or did not separate bloodstream from non-bloodstream data, as well as studies employing metagenomic approaches for resistance detection. Studies reporting insufficient or unclear data on antimicrobial resistance, whether genotypic or phenotypic, in cirrhosis patients were also excluded. Non-original research, including reviews, editorials, commentaries, conference abstracts, and case reports, were not considered for inclusion.

### Data extraction and quality assessment

Two authors (TP and KE) independently screened all retrieved articles and extracted data according to the predefined inclusion criteria. The quality of all included studies was assessed using the Newcastle-Ottawa Scale (NOS) for cohort and case-control studies and an adapted NOS for cross-sectional studies (Carra et al., 2025) (Supplementary File 1). The NOS evaluates risk of bias across selection, comparability, and outcome domains for cohort and case-control studies (maximum score 9), whereas the adapted NOS evaluates sample selection and outcome assessment for cross-sectional studies (maximum score 4). Studies were classified as low, moderate, or high risk of bias based on their total scores: 7–9 (cohort/case-control) or 3–4 (cross-sectional) for low, 4–6 or 2 for moderate, and 0–3 or 0–1 for high risk, respectively.

### Data analysis

Meta-analyses were conducted in R (version 4.5.2) using the meta and metafor packages. Pooled prevalence estimates of non-bloodstream infections, bacterial species, and antimicrobial-resistant phenotypes (including MDR, ESBL, MRSA, CRE, VRE, and fluoroquinolone resistance) were computed with 95% confidence intervals (CIs) using a random-effects model to account for between-study heterogeneity. When not directly reported, prevalence was calculated as the number of resistant isolates divided by the total tested. Heterogeneity was assessed using the I² statistic and interpreted as low (0–25%), moderate (25–75%), or high (>75%). Subgroup analyses were performed by publication period, study design, continent, infection source and type, patient age category, MELD score, and country income level. Countries were classified by income using the World Bank’s income group classification, which stratifies economies as low, lower-middle, upper-middle, or high income based on gross national income per capita via the World Bank Atlas method (World Bank, 2025). Study-level distributions of ARGs were tabulated and mapped across continents. Global distribution of resistance phenotypes and ARGs was visualized using R and QGIS to illustrate continental and regional patterns.

## Results

### Summary of included studies

A total of 31 studies comprising 3,162 non-bloodstream infections in patients with cirrhosis were included (Figure 1). SBP was the predominant infection type (n = 2,487; 78.7%), followed by colonization-type infections (n = 381; 12.1%) and urinary tract infections (n = 210; 6.6%). Less frequent infections included skin and soft tissue (n = 24; 0.8%), mixed (n = 22; 0.7%), respiratory tract (n = 15; 0.5%), and other infections (n = 23; 0.7%).

**Figure 2:**
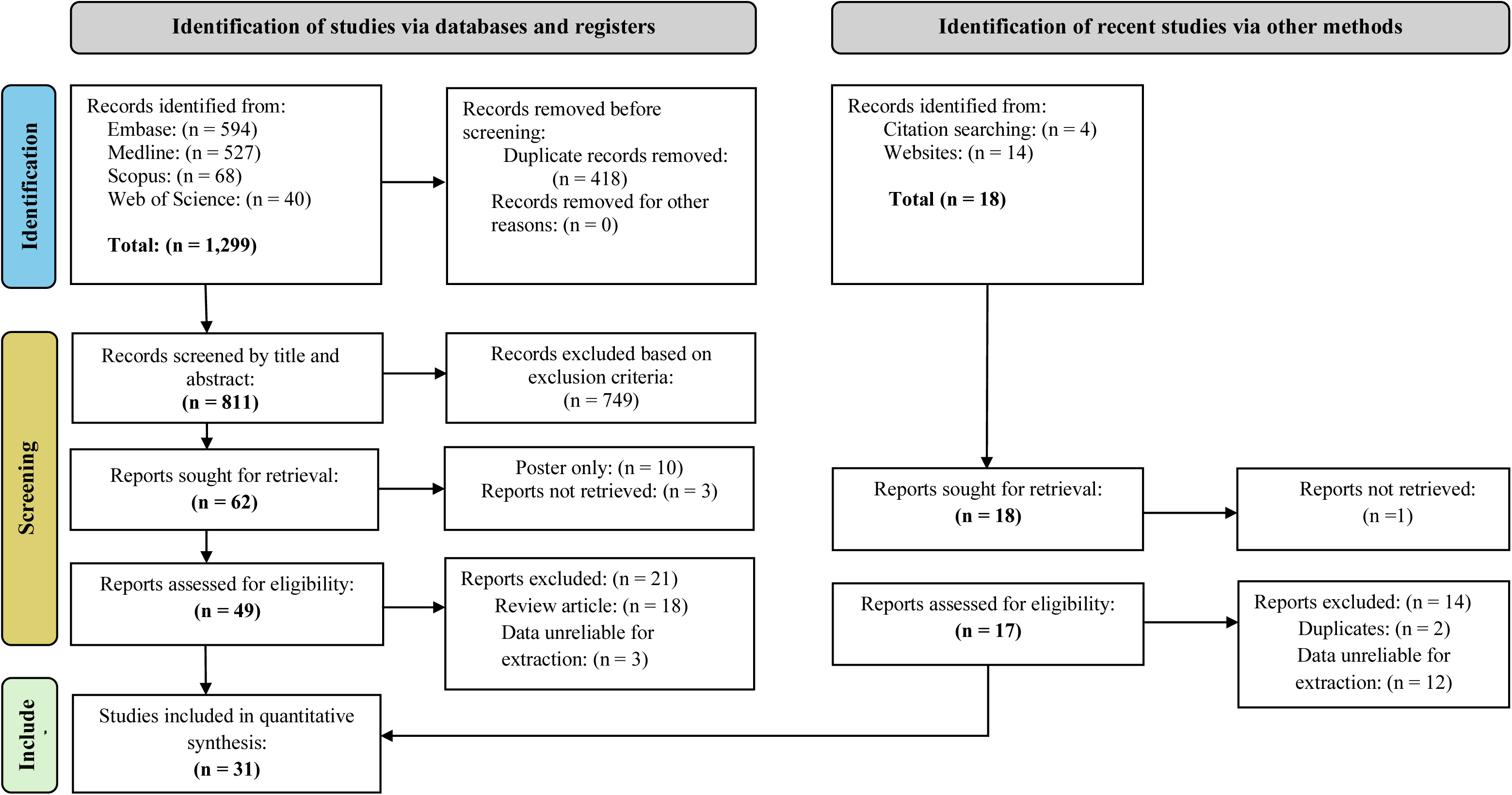
PRISMA flowchart of article selection progress.

The study selection process is shown in Figure 1. From 1,299 records identified through database searches, 418 duplicates were removed and 811 unique articles screened. After title and abstract review, 49 studies were assessed in full, of which 28 contained extractable data. Manual reference screening contributed 18 additional records, with 15 excluded, yielding a final total of 31 eligible studies for quantitative synthesis. A full list of the 31 included studies, along with their characteristics, is provided in Supplementary Table S1-3

The included studies spanned six continents, with the highest representation from Asia (n = 9) and Europe (n = 8), followed by Africa (n = 3), North America (n = 2), Oceania (n = 1), and South America (n = 1). Across the 27 studies reporting sex-specific data, male participants consistently predominated, accounting for more than half of all cases in every study (Supplementary Figure 2). Viral hepatitis dominated the aetiological profile of cirrhosis (unspecified hepatitis 57.9%; HBV 49.7%, HCV 11.9%), followed by alcohol-associated liver disease (35%), other causes (23.1%), and MASLD (3.7%). A summary of sex distribution (Fig. 2A), underlying aetiologies (Fig. 2B), and clinical characteristics reported across included studies (Fig. 2C) is shown in Figure 2

**Figure 2.**
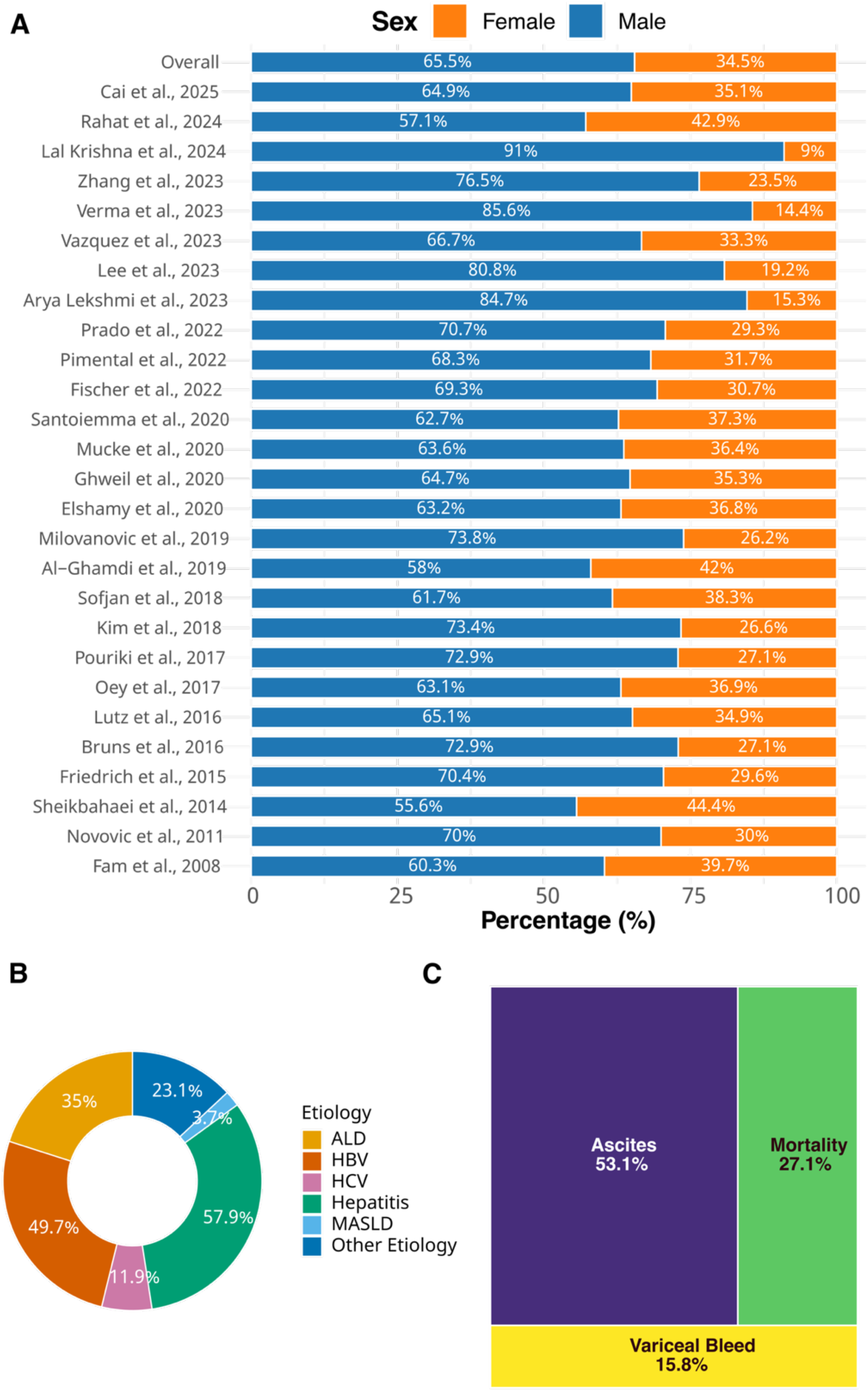
Demographic, aetiological, and clinical characteristics of cirrhosis-associated infections across studies. (A) Sex distribution of culture-positive infections in studies reporting sex-disaggregated data. (B) Aggregated aetiological profile of cirrhosis across studies. Etiology abbreviations: ALD, Alcohol-Associated Liver Disease; MASLD, Metabolic Dysfunction-Associated Steatotic Liver Disease; Hepatitis, hepatitis of unspecified type; HBV, Hepatitis B virus; HCV, Hepatitis C virus; Other Aetiology, other causes of cirrhosis. (C) Relative frequency of major clinical variables reported, shown as a treemap.

### Risk of bias assessments

Risk of bias was assessed using the Newcastle-Ottawa Scale for cohort and case-control studies and an adapted version for cross-sectional studies (Supplementary Figures 1–2, Supplementary File 1). Among 27 cohort studies, total scores ranged from 4.5 to 9, indicating generally moderate to high quality. Five studies scored the maximum 9, while most (n = 16, 59%) scored 7–8.5, with minor concerns mainly in participant selection or comparability. One study scored 4.5, reflecting moderate risk due to selection and comparability issues. The single case-control study scored 8.5, showing minor concerns in case selection and comparability. Among six cross-sectional studies, scores ranged from 2 to 4, with two studies showing low risk and the remainder moderate risk due to sampling or representativeness issues. Overall, no studies were deemed at high risk of bias, with most exhibiting low to moderate risk across domains.

### Non-bloodstream infection proportions across study characteristics

Pooled infection proportions varied across study characteristics (Table 1). Studies published between 2020–2025 showed a slightly higher pooled proportion of non-bloodstream infections (40%, 95% CI 31–51) compared with studies from 2008–2019 (35%, 95% CI 24–49). Across study designs, cross-sectional studies reported the highest pooled prevalence (53%, 95% CI 43–63), followed by prospective cohorts (47%, 95% CI 34–61) and retrospective analyses (29%, 95% CI 20–40).

**Table 1:** Pooled Proportions of Non-Bloodstream Infections in Cirrhosis Stratified by Study, Population, and Infection Characteristics.

| Variables | Included<br>Studies<br>(k) | Sample<br>Size (N) | Events<br>(n) | Pooled<br>Proportion<br>(95% CI) | I<sup>2</sup> (%) | p value<br>(heterogeneity) | p value<br>(subgroup<br>differences) |
| --- | --- | --- | --- | --- | --- | --- | --- |
| Publication Period |  |  |  |  |  |  |  |
| 2008–2019 | 9 | 2147 | 825 | 35(24 – 49) | 97.2 | <0.001 | 0.526 |
| 2020–2025 | 15 | 35202 | 10016 | 40(31 – 51) | 99.6 | <0.001 |  |
| Study design |  |  |  |  |  |  |  |
| Prospective Cohort | 9 | 1810 | 846 | 47(34 – 61) | 94.4 | <0.001 | <0.001 |
| Retrospective Cohort | 8 | 31897 | 9308 | 29(20 – 40) | 99.8 | <0.001 |  |
| Cross Sectional | 4 | 927 | 536 | 53(43 – 63) | 90 | <0.001 |  |
| Mixed | 2 | 427 | 101 | 24(20 – 28) | 0 | 0.558 |  |
| Case Control | 1 | 141 | 47 | 33(26 – 42) | NA | NA |  |
| Continent |  |  |  |  |  |  |  |
| Europe | 9 | 2170 | 772 | 36(26 – 48) | 94.5 | <0.001 | <0.001 |
| Asia | 8 | 19147 | 8369 | 47(34 – 60) | 96.6 | <0.001 |  |
| Africa | 3 | 535 | 190 | 38(24 – 54) | 95.1 | <0.001 |  |
| North America | 2 | 455 | 155 | 34(30 – 39) | 0 | 0.825 |  |
| Oceania | 1 | 12423 | 1096 | 9(8 – 9) | NA | NA |  |
| South America | 1 | 472 | 256 | 54(50 – 59) | NA | NA |  |
| Average Age (Years) |  |  |  |  |  |  |  |
| < 60 | 18 | 4387 | 1753 | 30(30 – 48) | 96.4 | <0.001 | 0.298 |
| ≥ 60 | 5 | 18242 | 7905 | 43(43 – 44) | 41.6 | 0.144 |  |
| MELD group |  |  |  |  |  |  |  |
| 19 | 8 | 19550 | 8268 | 32(22 – 44) | 96.6 | <0.001 | <0.001 |
| 20 - 24 | 3 | 836 | 404 | 49(44 – 54) | 48.9 | 0.141 |  |
| ≥25 | 5 | 755 | 340 | 50(34 – 66) | 94.6 | <0.001 |  |
| Infection Source |  |  |  |  |  |  |  |
| Community Acquired | 8 | 19347 | 12806 | 44(28 - 61) | 98.7 | <0.001 | 0.460 |
| Nosocomial | 8 | 19347 | 7247 | 35(21 - 53) | 96.9 | <0.001 |  |
| Type of Infection |  |  |  |  |  |  |  |
| SBP | 18 | 15855 | 2405 | 37(28 – 46) | 99.1 | <0.001 | <0.001 |
| UTI | 3 | 270 | 210 | 89(53 – 98) | 76.8 | 0.014 |  |
| Colonization | 3 | 782 | 399 | 57(37 – 75) | 95.2 | <0.001 |  |
| RTI | 1 | 63 | 15 | 24(14 – 36) | NA | NA |  |
| SSTI | 1 | 52 | 24 | 46(32 – 61) | NA | NA |  |
| Other | 1 | 54 | 23 | 43(29 – 57) | NA | NA |  |
| Mixed | 1 | 218 | 22 | 10(5 – 15) | NA | NA |  |

**Abbreviations:** k, number of studies in the subgroup; N, total sample size; n, number of infection events; CI, 95% confidence interval; I<sup>2</sup>, heterogeneity statistic; p (heterogeneity), Cochran’s Q test; p (subgroup), test for subgroup differences; SBP, spontaneous bacterial peritonitis; UTI, urinary tract infection; RTI, respiratory tract infection; SSTI, skin and soft tissue infection.

Geographic variation was evident, with South America reporting the highest pooled infection proportion (54%, 95% CI 50–59), followed by Asia (47%, 95% CI 34–60) and Africa (38%, 95% CI 24–54). Studies including populations aged ≥60 years showed a higher pooled infection proportion (43%, 95% CI 43–44) than those with younger cohorts (38%, 95% CI 30–48). Infection risk also increased with disease severity: patients with MELD ≥ 25 had a pooled proportion of 50% (95% CI 34–66), compared with 49% (95% CI 44–54) for MELD 20–24 and 30% (95% CI 22–44) for MELD < 19.

By infection source, community-acquired infections were more frequent (44%, 95% CI 28–61) than nosocomial infections (35%, 95% CI 21–53). Among specific infection types, urinary tract infections had the highest pooled prevalence (89%, 95% CI 53–98), followed by colonization-type infections (57%, 95% CI 37–75) and skin and soft tissue infections (46%, 95% CI 32–61).

### Pooled Proportions of Gram-negative and Gram-positive Bacteria

Among 10,324 cirrhosis patients with non-bacteraemic infections, Gram-negative bacteria accounted for 60% of infections (95% CI: 53–67), with study-specific estimates ranging from 24% to 92% (Figure 2). The most frequently reported Gram-negative pathogens were *Escherichia coli* (29%, 95% CI: 24–35) and *Klebsiella pneumoniae* (11%, 95% CI: 8–14), with *Pseudomonas aeruginosa* contributing 3% (95% CI: 2–5) of infections (Supplementary Figures 3–5). Gram-positive bacteria represented 39% of infections (95% CI: 32–45), with individual study estimates ranging from 8% to 72% (Figure 2; Supplementary Figures 6–8). The most commonly reported Gram-positive pathogens were Enterococci (14%, 95% CI: 10–20), *Staphylococcus aureus* (6%, 95% CI: 4–8), and *Enterococcus faecalis* (6%, 95% CI: 4–9) Study-level distributions of the 20 most frequently reported bacterial species are shown in Supplementary Figure 9A.

**Figure 3.**
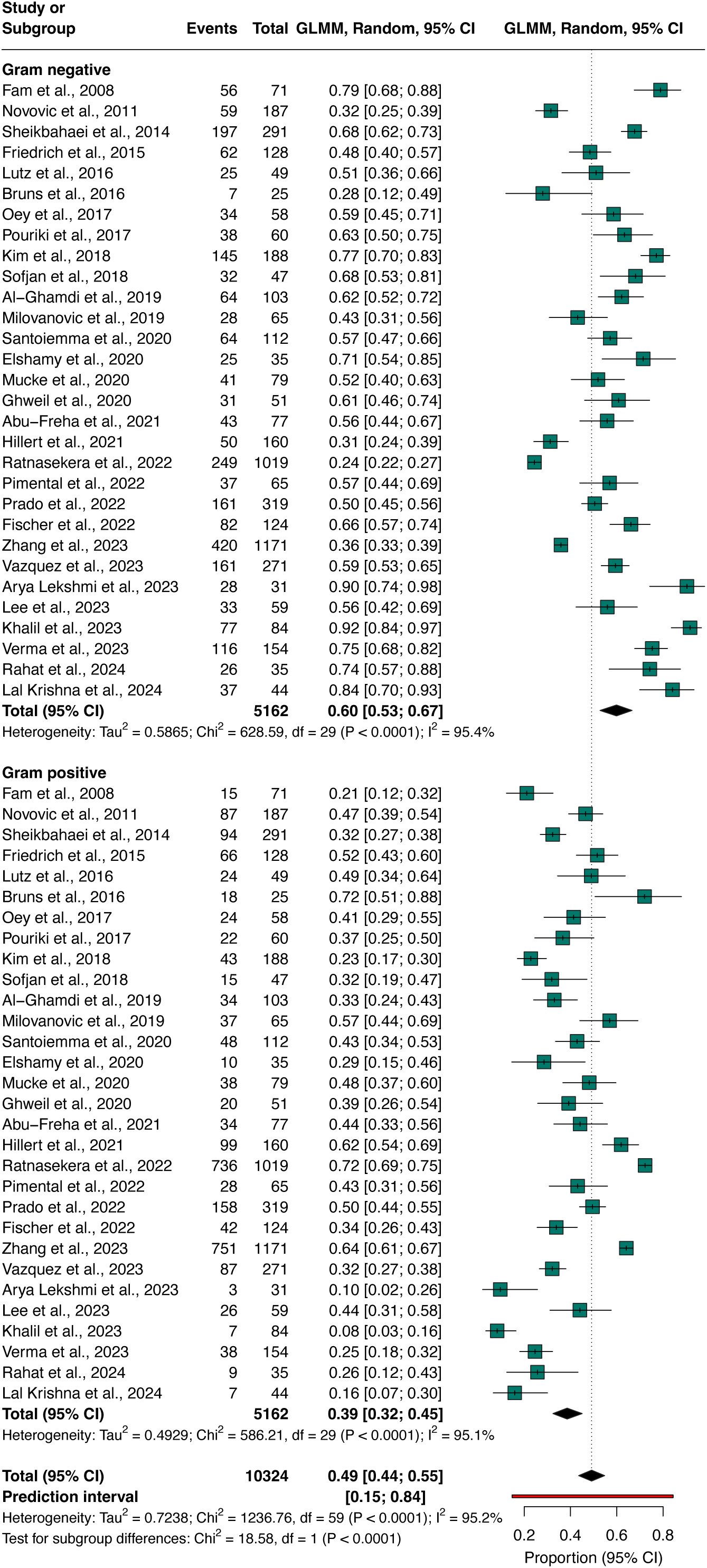
Pooled proportions of Gram-negative and Gram-positive bacteria in cirrhosis-associated non-bacteraemic infections. Forest plots display study-level prevalence (squares) with 95% confidence intervals, stratified by Gram status. Diamonds indicate pooled random-effects estimates with 95% confidence and prediction intervals.

### Distributions of Antimicrobial Resistance Profiles by infection type

Across cirrhosis-associated non-bloodstream infections, phenotypic resistance patterns varied markedly by pathogen, infection type, and period (Table 2). MRSA prevalence remained consistently low, with pooled proportions of 2–4% across SBP, mixed infections, and colonisation. In contrast, ESBL-producing Enterobacterales were substantially more common, particularly in UTI (25%) and colonisation isolates (32%), with overall ESBL prevalence reaching 11% and showing little temporal change. Species-specific estimates revealed ESBL *E. coli* at 7–24% depending on infection type, and ESBL *K. pneumoniae* remaining low in SBP (2–4%) but higher in colonisation (10%). VRE exhibited sharp stratification, ranging from 2–5% in SBP and mixed infections to 37% in colonisation samples. CRE remained rare overall (2%), although estimates were unstable due to high heterogeneity. Fluoroquinolone (QR) resistance was common in SBP (18%) and moderately high overall (16%), while ESC-R prevalence reached 13–22% across study periods. Multidrug resistance was pervasive, with MDR proportions of 18% in SBP, 19% in mixed infections, 54% in UTI, and up to 96% in colonisation, yielding an overall pooled MDR prevalence of 29% (Supplementary Table 1). Study-level distributions of major antimicrobial resistance phenotypes are shown in Supplementary Figure 9B.

**Table 2.** Pooled prevalence of antimicrobial-resistant phenotypes by country income level.

| Resistant Phenotype | Country income | No. of studies | Total isolates (n) | Pooled prevalence (95% CI) | *I*<sup>2</sup> | p value |
| --- | --- | --- | --- | --- | --- | --- |
| MRSA | High-income | 8 | 21,162 | 0.03 (0.02–0.05) | 85.7% | 0.45 |
|  | Upper-middle | 4 | 1,651 | 0.02 (0.01–0.04) | 82.5% |  |
|  | Lower-middle | 2 | 189 | 0.02 (0.01–0.06) | 0% |  |
| **Overall** | – | **14** | **23,002** | **0.02 (0.01–0.04)** | **87.0%** |  |
| ESBL | High-income | 12 | 2,453 | 0.10 (0.05–0.18) | 94.6% | 0.07 |
|  | Upper-middle | 3 | 1,360 | 0.10 (0.04–0.23) | 95.5% |  |
|  | Lower-middle | 3 | 260 | 0.24 (0.14–0.39) | 81.9% |  |
| **Overall** | – | **18** | **4,073** | **0.11 (0.07–0.18)** | **95.2%** |  |
| ESBL – E. coli | High-income | 11 | 2,182 | 0.08 (0.04–0.16) | 92.9% | <0.01 |
|  | Upper-middle | 2 | 1,295 | 0.04 (0.03–0.05) | 41.6% |  |
|  | Lower-middle | 2 | 106 | 0.14 (0.05–0.32) | 84.2% |  |
| **Overall** | – | **15** | **3,583** | **0.08 (0.05–0.14)** | **93.9%** |  |
| ESBL – K. pneumoniae | High-income | 6 | 625 | 0.04 (0.02–0.08) | 57.5% | 0.55 |
|  | Upper-middle | 2 | 1295 | 0.02 (0.00–0.08) | 94.1% |  |
|  | Lower-middle | 2 | 106 | 0.05 (0.02–0.11) | 0% |  |
| **Overall** | – | **10** | **2026** | **0.03 (0.02–0.05)** | **82.5%** |  |
| VRE | High-income | 11 | 21,348 | 0.03 (0.01–0.10) | 98.9% | <0.01 |
|  | Upper-middle | 4 | 1,651 | 0.03 (0.01–0.11) | 91.9% |  |
|  | Lower-middle | 2 | 198 | 0.21 (0.16–0.27) | 0% |  |
| **Overall** | – | **17** | **23,197** | **0.04 (0.02–0.09)** | **98.4%** |  |
| CRE | High-income | 3 | 19738 | 0.01 (0.01–0.01) | 0% | <0.01 |
|  | Upper-middle | 2 | 1,295 | 0.03 (0.00–0.28) | 98.1% |  |
|  | Lower-middle | 1 | 154 | 0.32 (0.25–0.40) | - |  |
| **Overall** | - | **6** | **21,187** | **0.02 (0.00–0.10)** | **98.9%** |  |
| QR | High-income | 4 | 381 | 0.20 (0.12–0.33) | 99.2% | <0.01 |
|  | Upper-middle | 1 | 1171 | 0.06 (0.05–0.08) | - |  |
| **Overall** | – | **5** | **1552** | **0.16 (0.08–0.28)** | **94.7%** |  |
| ESC-R | High-income | 6 | 19773 | 0.16 (0.08–0.30) | 90.8% | <0.01 |
|  | Upper-middle | 1 | 1,171 | 0.05 (0.04–0.06) | – |  |
| **Overall** | – | **7** | **20944** | **0.13 (0.06–0.25)** | **95.6%** |  |
| MDR | High-income | 14 | 21,661 | 0.22 (0.06–0.54) | 96.5% | 0.32 |
|  | Upper-middle | 3 | 1,360 | 0.41 (0.27–0.56) | 91.3% |  |
|  | Lower-middle | 4 | 298 | 0.47 (0.35–0.59) | 79.3% |  |
| **Overall** | – | **21** | **23,305** | **0.29 (0.14–0.50)** | **96.0%** |  |

No. of studies = included studies; Total isolates (n) = isolates analysed; Pooled prevalence (95% CI) = meta-analytic prevalence; *I*<sup>2</sup> = heterogeneity; p = test for subgroup differences. MRSA = methicillin-resistant *Staphylococcus aureus*; ESBL = extended-spectrum β-lactamase-producing Enterobacterales; VRE = vancomycin-resistant *Enterococcus* spp.; CRE = carbapenem-resistant Enterobacterales; QR = quinolone-resistant Gram-negative bacteria; ESC-R = extended-spectrum cephalosporin-resistant Gram-negative bacteria; MDR = multidrug-resistant (resistance to $\geq 3$ antimicrobial classes); – = not applicable.

### Disparities in AMR Across Country Income Levels

Across all phenotypes, we observed pronounced income-related disparities in antimicrobial resistance (Table 2). While MRSA prevalence was uniformly low across settings (2–3%), resistance among Gram-negative pathogens showed a clear gradient, with substantially higher burdens in lower–middle-income countries. ESBL-producing Enterobacterales were more than twice as prevalent in lower–middle-income countries (24%) compared with high- and upper–middle-income settings (both 10%), a pattern mirrored for *E. coli* (14% vs. 4–8%) and *K. pneumoniae*. Even starker differences were evident for high-priority threats: VRE increased from ∼3% in higher-income settings to 21% in lower–middle-income countries, and CRE rose from 1% to 32% across the same gradient. Multidrug resistance was widespread but consistently highest in lower–middle-income countries (47%), compared with 22–41% elsewhere.

### Global distribution of AMR phenotypes and genes

Across the six continents, ESBL-producing Enterobacterales were the most frequently reported AMR phenotypes. Europe reported the highest number of ESBL isolates (n = 170, 45%), followed by Asia (n = 142, 37%), South America (n = 22, 6%), and Africa (n = 19, 5%). In Europe and Asia, *E. coli* accounted for the majority of ESBL producers (66% and 56%, respectively), with *K. pneumoniae* contributing 25% and 8%. MRSA was detected globally at low frequencies (1–5 reports per continent), whereas VRE was concentrated in Europe (n = 216, 53%) and Asia (n = 75, 19%). CRE were largely restricted to Asia (n = 179, 45%). Oceania and North America reported only low numbers of MRSA and ESBL isolates.

Genotypic data from Africa, Asia, and Europe revealed 436 ARGs, with the highest counts in Asia (n = 251, 57%), followed by Europe (n = 93, 21%) and Africa (n = 65, 15%). Carbapenemase genes predominated (n = 209, 48%), followed by ESBL genes (n = 120, 28%). *bla*CTX-M was the most widespread ESBL determinant (Asia n = 44, 18%; Africa n = 6, 2%), while carbapenemases *bla*NDM (n = 37, 15%) and *bla*KPC (n = 24, 10%) were concentrated in Asia. *bla*VIM was largely reported in Europe (n = 18, 7%). Fluoroquinolone-resistance genes appeared only in Africa (n = 15, 6%), and *mecA* was restricted to Europe (n = 5, 2%). Vancomycin-resistance genes (*vanA*/*vanB*) were infrequently detected in Europe and Asia (<5%). These results reveal pronounced continental differences in both phenotypic and genotypic AMR distributions, with Asia exhibiting the greatest diversity and abundance of resistance determinants (Figure 4).

**Figure 4.**
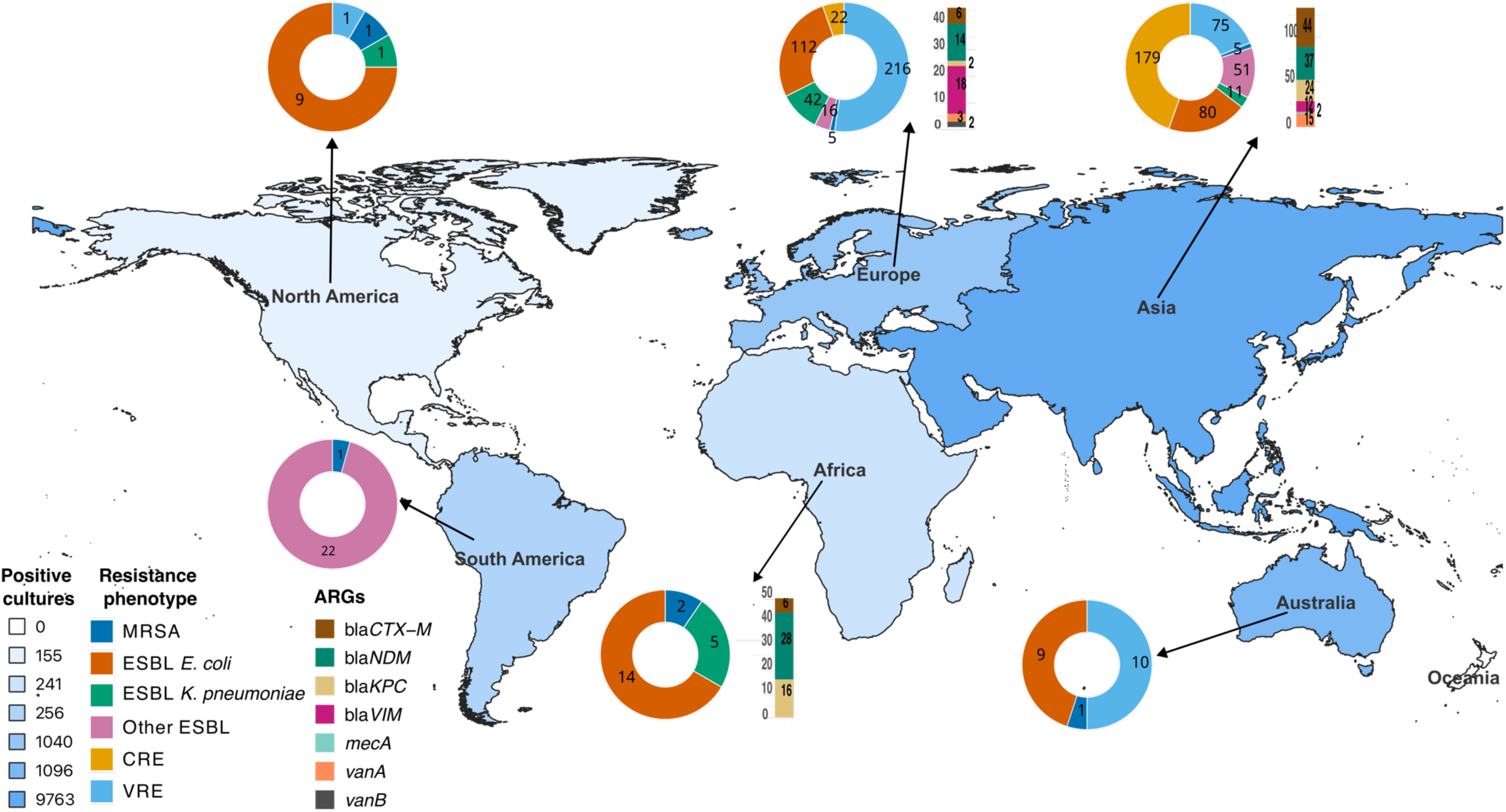
Global distribution of non-bloodstream infections, AMR types, and key resistance genes across continents. Donut charts showing the proportional distribution of major AMR phenotypes among clinical isolates across six continents. Categories include methicillin-resistant *Staphylococcus aureus* (MRSA), extended-spectrum β-lactamase (ESBL)-producing *Escherichia coli* and *Klebsiella pneumoniae*, other ESBL-producing species, carbapenem-resistant Enterobacterales (CRE), and vancomycin-resistant *Enterococcus* (VRE). Stacked bar plots illustrating the absolute counts of clinically important antimicrobial resistance genes (ARGs) detected across continents.

## Discussion

Our synthesis of 31 studies encompassing 3,162 non-bloodstream infections in cirrhosis indicates that SBP remains the predominant clinical presentation, reaffirming its central role among bacterial complications of ascites (Marciano et al., 2019; Tay et al., 2021). Considerable heterogeneity in infection profiles across regions, disease severities and patient groups likely reflects a combination of differing healthcare systems, surveillance practices, microbiological methods and host vulnerabilities (Paintsil et al., 2025; Piano et al., 2019; Y.-X. Tian et al., 2025). The higher prevalence observed in older adults and patients with advanced liver disease accords with established mechanisms—cirrhosis-associated immune dysfunction, impaired gut barrier integrity and increased bacterial translocation (Noor & Manoria, 2017). The greater infection proportions reported in more recent studies may reflect improved diagnostic vigilance and microbiological testing, evolving clinical practice, or genuine temporal changes in burden; these possibilities are not mutually exclusive and merit further study (Y.-X. Tian et al., 2025). Although community-acquired infections predominated overall (Ding et al., 2019), the unexpectedly high prevalence of urinary tract and colonization-type infections highlights that vulnerability extends beyond inpatient settings and underscores the need for broad surveillance and targeted prevention efforts across care settings (Bhattacharya et al., 2019).

Gram-negative bacteria dominate cirrhosis-associated non-bacteraemic infections, accounting for roughly 60 % of cases, with Escherichia coli and Klebsiella pneumoniae as the most frequently encountered pathogens. This predominance aligns with established pathophysiology: in cirrhosis, gut dysbiosis, increased intestinal permeability, and portosystemic shunting facilitate bacterial translocation from the gut lumen into ascitic fluid or other extra-intestinal sites, a recognized mechanism for spontaneous and non-spontaneous infections in advanced liver disease (Liu et al., 2025; Philips & Augustine, 2022; Ponziani et al., 2018; Tsiaoussis et al., 2015; Wozniak et al., 2024). The substantial contribution of Gram-positive bacteria, notably Enterococci and *Staphylococcus aureus* (Ding et al., 2019), suggests that infections in this population are frequently polymicrobial (Bunchorntavakul et al., 2016; Chen & To, 2024), which may reflect a combination of gut translocation, colonization of skin or mucosa, and exposure to healthcare environments (Bunchorntavakul et al., 2016; Pazhanivel et al., 2011; Raineri et al., 2022; Wozniak et al., 2024). The broad inter-study heterogeneity in pathogen distributions likely reflects genuine epidemiologic diversity but also variation in diagnostic protocols, sampling practices, and regional microbiota or antimicrobial pressure (Y.-X. Tian et al., 2025; Tozzo et al., 2022). These findings underscore the necessity for empiric antimicrobial therapy to be informed by local epidemiology, and support coverage for both Gram-negative and Gram-positive organisms — particularly in high-risk cirrhotic patients with ascites or immune compromise.

We also observed a substantial burden of antimicrobial resistance, with striking disparities by country income. MDR organisms, particularly ESBL-producing Enterobacterales, were commonly detected, especially in urinary tract and colonization isolates. This mirrors emerging data showing that cirrhosis-associated gut colonization often harbors resistant strains, which can later seed invasive infections (Hernández-Tejero et al., 2025; Verma et al., 2023; Wu et al., 2025). VRE and, alarmingly, CRE were more common in lower–middle-income settings and in colonization specimens, suggesting that structural determinants — such as limited microbiological diagnostics, overuse of broad-spectrum antibiotics, weak stewardship, and higher community transmission — shape global resistance patterns (Parra et al., 2025; F. Tian et al., 2023; Verma et al., 2023). The co-occurrence of multiple resistance phenotypes within the same patient groups, alongside the high prevalence of resistant colonization, suggests that cirrhotic patients may serve as reservoirs for onward transmission and are at substantial risk of subsequent difficult-to-treat infections (Kosuta et al., 2025; Prado et al., 2022; Verma et al., 2023). The steep resistance gradients, from high-resource to resource-limited settings, highlight growing inequities in AMR burden and underscore the urgent need for context-specific infection control, improved diagnostic capacity, equitable access to effective antimicrobials, and robust stewardship programs.

Pronounced geographical asymmetries in phenotypic and genotypic AMR profiles highlight the uneven global distribution of resistance selection pressures. Within our cohort of cirrhotic patients, ESBL-producing Enterobacterales—particularly *E. coli* and *K. pneumoniae* in Europe and Asia—were dominant, reflecting longstanding regional trends of third-generation cephalosporin use and established reservoirs of CTX-M–type ESBLs (Delvallez et al., 2025; Gales et al., 2023; Ramatla et al., 2023). VRE concentrations were similarly higher in Europe and Asia, while CRE and key carbapenemase genes (*blaNDM*, *blaKPC*) were almost exclusively detected in Asia, underscoring heterogeneity in antibiotic stewardship, infection-control capacity, and healthcare-associated transmission dynamics (Shrestha et al., 2021; Xia et al., 2024). Genotypic profiles mirrored these gradients: Asia harboured the highest diversity and abundance of ARGs, whereas Africa showed a more restricted but distinct resistance landscape, exemplified by fluoroquinolone-resistance genes not observed elsewhere. Limited detection of *mecA* and vancomycin-resistance operons may reflect either genuinely lower prevalence or, more likely, under-ascertainment due to sparse genomic surveillance in certain regions (Hendriksen et al., 2019; Totaro et al., 2025). Collectively, these continental disparities suggest deeply embedded structural determinants—including antimicrobial consumption patterns, diagnostic capacity, and regional mobility networks—that shape AMR evolution and circulation in cirrhotic populations (Furuya-Kanamori & Yakob, 2020; Mendelsohn et al., 2023), highlighting the urgent need for coordinated global genomic surveillance and context-specific interventions.

### Limitations

This meta-analysis is constrained by heterogeneity in study design, populations, microbiological methods, and reporting, resulting in high between-study variability. Geographic coverage is uneven, with limited data from many low- and middle-income regions, which may bias global AMR patterns. Genotypic surveillance is largely restricted to high-income settings, so the low detection of resistance determinants may reflect under-ascertainment rather than true absence. Finally, reliance on observational and published studies introduces potential selection and publication bias and may underrepresent mild or community-acquired infections, limiting generalizability to all cirrhotic populations.

### Conclusion

Our synthesis reveals pronounced disparities in the prevalence and antimicrobial resistance of cirrhosis-associated non-bloodstream infections, with particularly high multidrug resistance in lower–middle-income countries. These findings emphasize the need for integrated phenotypic and genomic surveillance in resource-limited settings to guide empiric therapy and mitigate the spread of resistant pathogens, ultimately improving outcomes for patients with cirrhosis.

## DATA AVAILABILITY STATEMENT

All data generated or analysed during this study are included in this published article and its Supplementary Material. Further inquiries can be directed to the corresponding author.

## AUTHOR CONTRIBUTIONS

E.K.P. and D.L.S.: conceptualization. E.K.P.: formal analysis. D.L.S. and J.J.W.: supervision. C.O., K.E., and P.T., validation. E.K.P. C.O., K.E., and P.T: writing—original draft. All authors (E.K.P., C.O., K.E., and P.T J.J.P., and D.L.S.) contributed to writing—review and editing, and all authors read and gave final approval of the version to be submitted.

