## Supplemental Figure 1 for "Burden and Genomic Landscape of Antimicrobial Resistance in Non-Bloodstream Infections Among Patients with Cirrhosis: A Systematic Review and Meta-Analysis"

### Supplementary File 1

### PRISMA Checklist

| Section and Topic | Item # | Checklist item | Location where item is reported |
|---|---|---|---|
| **TITLE** | | | |
| Title | 1 | Identify the report as a systematic review. | 1 - Title |
| **ABSTRACT** | | | |
| Abstract | 2 | See the PRISMA 2020 for Abstracts checklist. | |
| **INTRODUCTION** | | | |
| Rationale | 3 | Describe the rationale for the review in the context of existing knowledge. | 1-2 – Introduction, paragraph 2-3 |
| Objectives | 4 | Provide an explicit statement of the objective(s) or question(s) the review addresses. | 2 – Introduction, paragraph 4 |
| **METHODS** | | | |
| Eligibility criteria | 5 | Specify the inclusion and exclusion criteria for the review and how studies were grouped for the syntheses. | 3 – Methods, paragraph 4-5 |
| Information sources | 6 | Specify all databases, registers, websites, organisations, reference lists and other sources searched or consulted to identify studies. Specify the date when each source was last searched or consulted. | 2 – Methods, paragraph 2 |
| Search strategy | 7 | Present the full search strategies for all databases, registers and websites, including any filters and limits used. | 2 – Methods, paragraph 2 |
| Selection process | 8 | Specify the methods used to decide whether a study met the inclusion criteria of the review, including how many reviewers screened each record and each report retrieved, whether they worked independently, and if applicable, details of automation tools used in the process. | 2 – 3 Methods, paragraph 3 |
| Data collection process | 9 | Specify the methods used to collect data from reports, including how many reviewers collected data from each report, whether they worked independently, any processes for obtaining or confirming data from study investigators, and if applicable, details of automation tools used in the process. | 3 – Methods, paragraph 6 |
| Data items | 10a | List and define all outcomes for which data were sought. Specify whether all results that were compatible with each outcome domain in each study were sought (e.g. for all measures, time points, analyses), and if not, the methods used to decide which results to collect. | |

| Section and Topic | Item # | Checklist item | Location where item is reported |
|---|---|---|---|
| | 10b | List and define all other variables for which data were sought (e.g. participant and intervention characteristics, funding sources). Describe any assumptions made about any missing or unclear information. | |
| Study risk of bias assessment | 11 | Specify the methods used to assess risk of bias in the included studies, including details of the tool(s) used, how many reviewers assessed each study and whether they worked independently, and if applicable, details of automation tools used in the process. | 3 – Methods, paragraph 6 |
| Effect measures | 12 | Specify for each outcome the effect measure(s) (e.g. risk ratio, mean difference) used in the synthesis or presentation of results. | 4 – Methods, paragraph 7 |
| Synthesis methods | 13a | Describe the processes used to decide which studies were eligible for each synthesis (e.g. tabulating the study intervention characteristics and comparing against the planned groups for each synthesis (item #5)). | 4 – Methods, paragraph 7 |
| | 13b | Describe any methods required to prepare the data for presentation or synthesis, such as handling of missing summary statistics, or data conversions. | 4 – Methods, paragraph 7 |
| | 13c | Describe any methods used to tabulate or visually display results of individual studies and syntheses. | 4 – Methods, paragraph 7 |
| | 13d | Describe any methods used to synthesize results and provide a rationale for the choice(s). If meta-analysis was performed, describe the model(s), method(s) to identify the presence and extent of statistical heterogeneity, and software package(s) used. | 4 – Methods, paragraph 7 |
| | 13e | Describe any methods used to explore possible causes of heterogeneity among study results (e.g. subgroup analysis, meta-regression). | 4 – Methods, paragraph 7 |
| | 13f | Describe any sensitivity analyses conducted to assess robustness of the synthesized results. | 4 – Methods, paragraph 7 |
| Reporting bias assessment | 14 | Describe any methods used to assess risk of bias due to missing results in a synthesis (arising from reporting biases). | 3 & 4 - Methods, paragraph 6 & 7 |
| Certainty assessment | 15 | Describe any methods used to assess certainty (or confidence) in the body of evidence for an outcome. | 4 – Methods, paragraph 7 |
| **RESULTS** | | | |

| Section and Topic | Item # | Checklist item | Location where item is reported |
|---|---|---|---|
| Study selection | 16a | Describe the results of the search and selection process, from the number of records identified in the search to the number of studies included in the review, ideally using a flow diagram. | 4 & 5 – Results paragraph 2, Figure 1 |
| | 16b | Cite studies that might appear to meet the inclusion criteria, but which were excluded, and explain why they were excluded. | 4 – Results, paragraph 2 |
| Study characteristics | 17 | Cite each included study and present its characteristics. | Table 1 |
| Risk of bias in studies | 18 | Present assessments of risk of bias for each included study. | |
| Results of individual studies | 19 | For all outcomes, present, for each study: (a) summary statistics for each group (where appropriate) and (b) an effect estimate and its precision (e.g. confidence/credible interval), ideally using structured tables or plots. | Table 1 & 2, Figure 2 |
| Results of syntheses | 20a | For each synthesis, briefly summarise the characteristics and risk of bias among contributing studies. | |
| | 20b | Present results of all statistical syntheses conducted. If meta-analysis was done, present for each the summary estimate and its precision (e.g. confidence/credible interval) and measures of statistical heterogeneity. If comparing groups, describe the direction of the effect. | 5 – 13, Methods , paragraph 1 - 11 |
| | 20c | Present results of all investigations of possible causes of heterogeneity among study results. | 5 – 13, Methods , paragraph 1 – 11 |
| | 20d | Present results of all sensitivity analyses conducted to assess the robustness of the synthesized results. | 5 – 13, Methods , paragraph 1 – 11 |
| Reporting biases | 21 | Present assessments of risk of bias due to missing results (arising from reporting biases) for each synthesis assessed. | 5 – 13, Methods , paragraph 1 – 11 |
| Certainty of evidence | 22 | Present assessments of certainty (or confidence) in the body of evidence for each outcome assessed. | 5 – 13, Methods , paragraph 1 - 11 |
| **DISCUSSION** | | | |

| Section and Topic | Item # | Checklist item | Location where item is reported |
|---|---|---|---|
| Discussion | 23a | Provide a general interpretation of the results in the context of other evidence. | 13 – Discussion, Paragraph1 |
| | 23b | Discuss any limitations of the evidence included in the review. | 15 – Discussion, Paragraph 6 & 7 |
| | 23c | Discuss any limitations of the review processes used. | 15 – Discussion, Paragraph 7 |
| | 23d | Discuss implications of the results for practice, policy, and future research. | 13 – 15 Discussion, paragraph 1 - 5 |
| **OTHER INFORMATION** | | | |
| Registration and protocol | 24a | Provide registration information for the review, including register name and registration number, or state that the review was not registered. | |
| | 24b | Indicate where the review protocol can be accessed, or state that a protocol was not prepared. | |
| | 24c | Describe and explain any amendments to information provided at registration or in the protocol. | |
| Support | 25 | Describe sources of financial or non-financial support for the review, and the role of the funders or sponsors in the review. | |
| Competing interests | 26 | Declare any competing interests of review authors. | |
| Availability of data, code and other materials | 27 | Report which of the following are publicly available and where they can be found: template data collection forms; data extracted from included studies; data used for all analyses; analytic code; any other materials used in the review. | |

*From:* Page MJ, McKenzie JE, Bossuyt PM, Boutron I, Hoffmann TC, Mulrow CD, et al. The PRISMA 2020 statement: an updated guideline for reporting systematic reviews. BMJ 2021;372:n71. doi: 10.1136/bmj.n71. This work is licensed under CC BY 4.0. To view a copy of this license, visit https://creativecommons.org/licenses/by/4.0/

### Search Strategy

**Conducted on ……………**

**Database: Medline (via PubMed or Ovid)**

| # | *Searches* | Results |
|---|---|---|
| **1** | ("Cirrhosis"[Mesh] OR "Liver Diseases"[Mesh] OR cirrhosis[tiab] OR "liver disease"[tiab] OR "chronic liver disease"[tiab] OR "end-stage liver disease"[tiab]) | |
| **2** | ("antimicrobial resistance gene*"[tw] OR "antibiotic resistance gene*"[tw] OR "AMR gene*"[tw] OR ARG*[tw] OR "phenotypic resistance"[tw] OR "resistance phenotype*"[tw] OR "antimicrobial resist*"[tw] OR "antibiotic resist*"[tw] OR "multidrug resist*"[tw] OR "resistant bacteria"[tw] OR "AMR"[tw]) | |
| **3** | ("non-bloodstream infection"[tw] OR "non bloodstream infection"[tw] OR "ascitic fluid"[tw] OR "spontaneous bacterial peritonitis"[tw] OR SBP[tw] OR "urinary tract infection"[tw] OR UTI[tw] OR pneumonia[tw] OR "respiratory infection"[tw] OR "wound infection"[tw] OR "skin infection"[tw] OR "soft tissue infection"[tw] OR "bacterial infection"[tw]) | |
| **4** | **#1 AND #2 AND #3** | **527** |

**Database: Embase (via Ovid )**

| # | *Searches* | Results |
|---|---|---|
| **1** | 'cirrhosis'/exp OR 'liver disease'/exp OR cirrhosis:ti,ab OR 'liver disease':ti,ab OR 'chronic liver disease':ti,ab OR 'end-stage liver disease':ti,ab | |
| **2** | 'antimicrobial resistance gene*':ti,ab OR 'antibiotic resistance gene*':ti,ab OR 'AMR gene*':ti,ab OR ARG*:ti,ab OR 'phenotypic resistance':ti,ab OR 'resistance phenotype*':ti,ab OR 'antimicrobial resist*':ti,ab OR 'antibiotic resist*':ti,ab OR 'multidrug resist*':ti,ab OR 'resistant bacteria':ti,ab OR 'AMR':ti,ab | |
| **3** | 'non-bloodstream infection':ti,ab OR 'non bloodstream infection':ti,ab OR 'ascitic fluid':ti,ab OR 'spontaneous bacterial peritonitis':ti,ab OR SBP:ti,ab OR 'urinary tract infection':ti,ab OR UTI:ti,ab OR pneumonia:ti,ab OR 'respiratory infection':ti,ab OR 'wound infection':ti,ab OR 'skin infection':ti,ab OR 'soft tissue infection':ti,ab OR 'bacterial infection':ti,ab | |
| **3** | **#1 AND #2 AND #3** | **594** |

**Database: ISI Web of Science**

| # | *Searches* | *Results* |
|---|---|---|

| | | |
|---|---|---|
| **1** | TS=("cirrhosis" OR "liver disease" OR "chronic liver disease" OR "end-stage liver disease") | |
| **2** | TS=("antimicrobial resistance gene*" OR "antibiotic resistance gene*" OR "AMR gene*" OR ARG* OR "phenotypic resistance" OR "resistance phenotype*" OR "antimicrobial resist*" OR "antibiotic resist*" OR "multidrug resist*" OR "resistant bacteria" OR "AMR") | |
| **3** | TS=("non-bloodstream infection" OR "non bloodstream infection" OR "ascitic fluid" OR "spontaneous bacterial peritonitis" OR SBP OR "urinary tract infection" OR UTI OR pneumonia OR "respiratory infection" OR "wound infection" OR "skin infection" OR "soft tissue infection" OR "bacterial infection") | |
| **3** | **1 AND #2 AND #3** | **40** |

**Database: Scopus**

| # | ***Searches*** | ***Results*** |
|---|---|---|
| **1** | TITLE-ABS("cirrhosis" OR "liver disease" OR "chronic liver disease" OR "end-stage liver disease") AND TITLE-ABS("antimicrobial resistance gene*" OR "antibiotic resistance gene*" OR "AMR gene*" OR ARG* OR "phenotypic resistance" OR "resistance phenotype*" OR "antimicrobial resist*" OR "antibiotic resist*" OR "multidrug resist*" OR "resistant bacteria" OR "AMR") AND TITLE-ABS("non-bloodstream infection" OR "non bloodstream infection" OR "ascitic fluid" OR "spontaneous bacterial peritonitis" OR SBP OR "urinary tract infection" OR UTI OR pneumonia OR "respiratory infection" OR "wound infection" OR "skin infection" OR "soft tissue infection" OR "bacterial infection") | |
| | **Total** | **68** |

### NEWCASTLE-OTTAWA QUALITY ASSESSMENT SCALE (NOS)

This scale is used to assess the quality of nonrandomized studies, including case-control and cohort studies, in systematic reviews and meta-analyses.

**Outcome Categories**

Each question is rated as follows:

**"Yes"** = 1 point (high quality)

**"Somewhat"** = 0.5 points (moderate quality)

**"No"** = 0 points (low quality)

---

**Cohort Study**

**Selection Criteria**

1. **Representativeness of the Exposed Cohort**

   a) Truly representative of the average population in the community. (**Yes =**

   b) Somewhat representative of the average population in the community. (**Somewhat = 0.5**)

   c) Selected group of users (e.g., nurses, volunteers). (**Somewhat = 0.5**)

   d) No description of cohort derivation. (**No = 0**)

2. **Selection of the Non-Exposed Cohort**

   a) Drawn from the same community as the exposed cohort. (**Yes = 1**)

   b) Drawn from a different source. (**Somewhat = 0.5**)

   c) No description of derivation of non-exposed cohort. (**No = 0**)

3. **Ascertainment of Exposure**

   a) Secure record (e.g., surgical records). (**Yes = 1**)

   b) Structured interview. (**Yes = 1**)

   c) Written self-report. (**Somewhat = 0.5**)

   d) No description. (**No = 0**)

4. **Demonstration That Outcome Was Not Present at Start of Study**

   a) Yes. (**Yes = 1**)

   b) Somewhat. (**Somewhat = 0.5**)

   c) No. (**No = 0**)

**Comparability Criteria**

1. **Control for Most Important Factor**

   a) Yes. (**Yes = 1**)

   b) Somewhat. (**Somewhat = 0.5**)

   c) No. (**No = 0**)

2. **Control for Additional Factor**

a) Yes. (**Yes = 1**)

b) Somewhat. (**Somewhat = 0.5**)

c) No. (**No = 0**)

**Outcome Criteria**

1. **Assessment of Outcome**

   a) Independent blind assessment. (**Yes = 1**)

   b) Record linkage. (**Yes = 1**)

   c) Self-report. (**Somewhat = 0.5**)

   d) No description. (**No = 0**)

2. **Sufficiency of Follow-Up Duration**

   a) Yes. (**Yes = 1**)

   b) No. (**No = 0**)

   c) Somewhat. (**Somewhat = 0.5**)

3. **Adequacy of Follow-Up of Cohorts**

   a) Complete follow-up – all subjects accounted for. (**Yes = 1**)

   b) Subjects lost to follow-up unlikely to introduce bias (small number lost, > X%, or description provided of those lost). (**Somewhat = 0.5**)

   c) Follow-up rate < X% and no description of those lost. (**Somewhat = 0.5**)

   d) No statement. (**No = 0**)

---

**Case-Control Study**

**Selection Criteria**

1. **Adequacy of Case Definition**

   a) Yes, with independent validation. (**Yes = 1**)

   b) Yes, e.g., record linkage or self-reports. (**Somewhat = 0.5**)

   c) No description. (**No = 0**)

2. **Representativeness of the Cases**

   a) Consecutive or obviously representative series of cases. (**Yes = 1**)

   b) Somewhat representative, potential selection biases, or not stated. (**Somewhat = 0.5**)

   c) No description. (**No = 0**)

3. **Selection of Controls**

   a) Community controls. (**Yes = 1**)

   b) Hospital controls. (**Somewhat = 0.5**)

   c) No description. (**No = 0**)

4. **Definition of Controls**

a) No history of disease (endpoint). (**Yes** = **1**)

b) Somewhat described source. (**Somewhat** = **0.5**)

c) No description of source. (**No** = **0**)

**Comparability of Cases and Controls**

1. **Control for Most Important Factor**

a) Yes. (**Yes** = **1**)

b) Somewhat. (**Somewhat** = **0.5**)

c) No. (**No** = **0**)

2. **Control for Additional Factor**

a) Yes. (**Yes** = **1**)

b) Somewhat. (**Somewhat** = **0.5**)

c) No. (**No** = **0**)

**Exposure Criteria**

1. **Ascertainment of Exposure**

a) Secure record (e.g., surgical records). (**Yes** = **1**)

b) Structured interview, blind to case/control status. (**Yes** = **1**)

c) Interview not blinded to case/control status. (**Somewhat** = **0.5**)

d) Written self-report or medical record only. (**Somewhat** = **0.5**)

e) No description. (**No** = **0**)

2. **Consistency of Exposure Measurement for Cases and Controls**

a) Yes. (**Yes** = **1**)

b) Somewhat. (**Somewhat** = **0.5**)

c) No. (**No** = **0**)

3. **Non-Response Rate**

a) Same rate for both groups. (**Yes** = **1**)

b) Non-respondents described. (**Somewhat** = **0.5**)

c) Rate different with no description. (**No** = **0**)

#### Cross-Sectional Studies

**NOS-xs2: Adaptation of the Newcastle-Ottawa Scale Simplified Version for Descriptive Cross-Sectional Studies (Prevalence Studies)**

**Total stars: 4**
**Risk of Bias (RoB) categories:**

- Low RoB: 3–4 stars
- Moderate RoB: 2 stars
- High RoB: 0–1 star

**Domain 1: Study Sample Selection (Maximum 2 stars)**

**Item 1: Representativeness of the Sample (1 star)**

- a) Truly representative of the target population → 1 star
- b) Somewhat representative → 1 star (context-dependent)
- c) Convenience sample / not representative → 0 star

**Item 2: Sample Size (1 star)**

- a) Justified and sufficient (a priori or post hoc power) → 1 star
- b) Not justified / insufficient → 0 star

---

**Domain 2: Assessment of Outcome(s) (Maximum 2 stars)**

**Item 3: Assessment of Outcome (2 stars)**

- a) Gold standard method for outcome → 2 stars
- b) Acceptable / validated alternative method → 1 star
- c) Poor or unvalidated method / no description → 0 star

#### Supplementary Table:

**Table 1: Phenotypic antimicrobial resistance prevalence in bacterial pathogens from cirrhosis-associated non-bloodstream infections, stratified by infection type and study period.**

| Variable | No. of studies | Total isolates | Pooled Proportion (95% CI) | $I^2$ (%) | p value (heterogeneity) | p value (subgroup differences) |
|---|---|---|---|---|---|---|
| **MRSA** | | | | | | |
| SBP | 8 | 2874 | 2 (1 – 4) | 74 | <0.01 | **0.41** |
| UTI | 1 | 65 | 3 (0 – 11) | NA | NA | |
| Mixed Infection | 4 | 19909 | 4 (2 – 7) | 70 | 0.02 | |
| Colonisation | 1 | 154 | 2 (0 – 6) | NA | NA | |
| 2002–2019 | 5 | 649 | 3 (2 – 6) | 28 | 0.23 | **0.44** |
| 2020–2025 | 9 | 22353 | 2 (1 – 4) | 91 | <0.01 | |
| **Overall** | **14** | **23002** | **2 (1 – 4)** | **87** | **<0.01** | |
| **ESBL** | | | | | | |
| SBP | 12 | 3080 | 8 (4 – 15) | 92 | <0.01 | **<0.01** |
| UTI | 1 | 65 | 25 (15 – 37) | NA | NA | |
| Mixed Infection | 2 | 395 | 9 (7 – 12) | 2 | 0.31 | |
| Colonisation | 3 | 533 | 32 (28 – 36) | 0 | 0.45 | |
| 2002-2019 | 8 | 779 | 11 (5 – 20) | 84 | <0.01 | **0.77** |
| 2020-2025 | 10 | 3294 | 12 (6 – 23) | 97 | <0.01 | |
| **Overall** | **18** | **4073** | **11 (7 – 18)** | **95** | **<0.01** | |
| **ESBL *E. coli*** | | | | | | |
| SBP | 12 | 3080 | 7 (4 – 13) | 91 | <0.01 | **<0.01** |
| UTI | - | NA | NA | NA | NA | |
| Mixed Infection | 1 | 124 | 6 (2 – 11) | NA | NA | |
| Colonisation | 2 | 379 | 24 (20 – 28) | 0 | 0.80 | |
| 2002–2019 | 7 | 714 | 8 (4 – 17) | 84 | <0.01 | **0.99** |
| 2020–2025 | 8 | 2869 | 8 (4 – 17) | 96 | <0.01 | |
| **Overall** | **15** | **3583** | **8 (5 – 14)** | **93** | **<0.01** | |
| **ESBL *K. pneumoniae*** | | | | | | |
| SBP | 8 | 1583 | 2 (1 – 4) | 54 | 0.03 | **<0.01** |
| UTI | NA | NA | NA | NA | NA | |
| Mixed Infection | 1 | 124 | 6 (2 – 11) | NA | NA | |
| Colonisation | 1 | 319 | 10 (7 – 14) | NA | NA | |
| 2002-2019 | 3 | 176 | 3 (1 – 7) | 0 | 0.67 | **0.61** |
| 2020-2025 | 7 | 1850 | 3 (2 – 7) | 87 | <0.01 | |
| **Overall** | **10** | **2026** | **3 (2 – 5)** | **82** | **<0.01** | |
| **VRE** | | | | | | |
| SBP | 9 | 2882 | 2 (1– 5) | 84 | <0.01 | **<0.01** |
| UTI | 1 | 65 | 11 (4 – 21) | NA | NA | |
| Mixed Infection | 4 | 19717 | 2 (0 – 10) | 98 | <0.01 | |
| Colonisation | 3 | 533 | 37 (24 – 51) | 93 | <0.01 | |
| 2002-2019 | 6 | 649 | 6 (2 – 15) | 92 | <0.01 | **0.49** |

| | | | | | | |
|---|---|---|---|---|---|---|
| 2020-2025 | 11 | 22548 | 3 (1 – 10) | 98 | <0.01 | |
| **Overall** | **17** | **23197** | **4 (2 – 9)** | **98** | **<0.01** | |
| **CRE** | | | | | | |
| SBP | 2 | 1236 | 0 (0 – 1) | 27 | <0.24 | **0.12** |
| UTI | NA | NA | NA | NA | NA | |
| Mixed Infection | 2 | 19478 | 3 (0 – 25) | 99 | <0.01 | |
| Colonisation | 2 | 473 | 5 (0 – 56) | 97 | <0.01 | |
| 2002-2019 | NA | NA | NA | NA | NA | **NA** |
| 2020-2025 | 6 | 21187 | 2 (0 – 10) | 99 | <0.01 | |
| **Overall** | **6** | **21187** | **2 (0 – 10)** | **99** | **<0.01** | |
| **QR** | | | | | | |
| SBP | 4 | 1473 | 18 (9-34) | 96 | <0.01 | **0.26** |
| UTI | NA | NA | NA | NA | NA | |
| Mixed Infection | 1 | 79 | 10 (4 – 19) | NA | NA | |
| Colonisation | NA | NA | NA | NA | NA | |
| 2002-2019 | 2 | 237 | 21 (10 – 39) | 91 | <0.01 | **0.33** |
| 2020-2025 | 4 | 20669 | 16 (7 – 24) | 96 | <0.01 | |
| **Overall** | **6** | **20906** | **16 (8 – 25)** | **94** | **<0.01** | |
| **ESC-R** | | | | | | |
| SBP | 6 | 1590 | 13 (5 – 27) | 96 | <0.01 | **0.69** |
| UTI | NA | NA | NA | NA | NA | |
| Mixed Infection | 1 | 19354 | 15 (15 – 16) | NA | NA | |
| Colonisation | NA | NA | NA | NA | NA | |
| 2002-2019 | 3 | 295 | 9 (2 – 26) | 92 | <0.01 | **0.20** |
| 2020-2025 | 4 | 20649 | 22 (9 – 42) | 99 | <0.01 | |
| **Overall** | **7** | **20944** | **15 (7 – 30)** | **98** | **<0.01** | |
| **MDR** | | | | | | |
| SBP | 13 | 2990 | 18 (10 – 31) | 97 | <0.01 | **<0.01** |
| UTI | 1 | 65 | 54 (41– 66) | NA | NA | |
| Mixed Infection | 4 | 19717 | 19 (7 – 40) | 89 | <0.01 | |
| Colonisation | 3 | 533 | 96 (24 – 100) | 70 | 0.03 | |
| 2002-2019 | 7 | 594 | 22 (7 – 53) | 93 | <0.01 | **0.59** |
| 2020-2025 | 13 | 3357 | 33 (12 – 64) | 97 | <0.01 | |
| **Overall** | **21** | **23305** | **29 (14 – 50)** | **96** | **<0.01** | |

**Abbreviations:** CI: confidence interval; $I^2$: heterogeneity statistic; p (heterogeneity): Cochran's Q test for within-subgroup heterogeneity; p (subgroup differences): test for differences between subgroups; SBP: spontaneous bacterial peritonitis; UTI: urinary tract infection; MRSA: methicillin-resistant *Staphylococcus aureus*; ESBL: extended-spectrum β-lactamase; ESBL E. coli: ESBL-producing *Escherichia coli*; ESBL K. pneumoniae: ESBL-producing *Klebsiella pneumoniae*; VRE: vancomycin-resistant *Enterococcus*; CRE: carbapenem-resistant *Enterobacterales*; QR: quinolone-resistant isolates; ESC-R: third-generation cephalosporin–resistant isolates; MDR: multidrug-resistant.

**Supplementary Figures:**

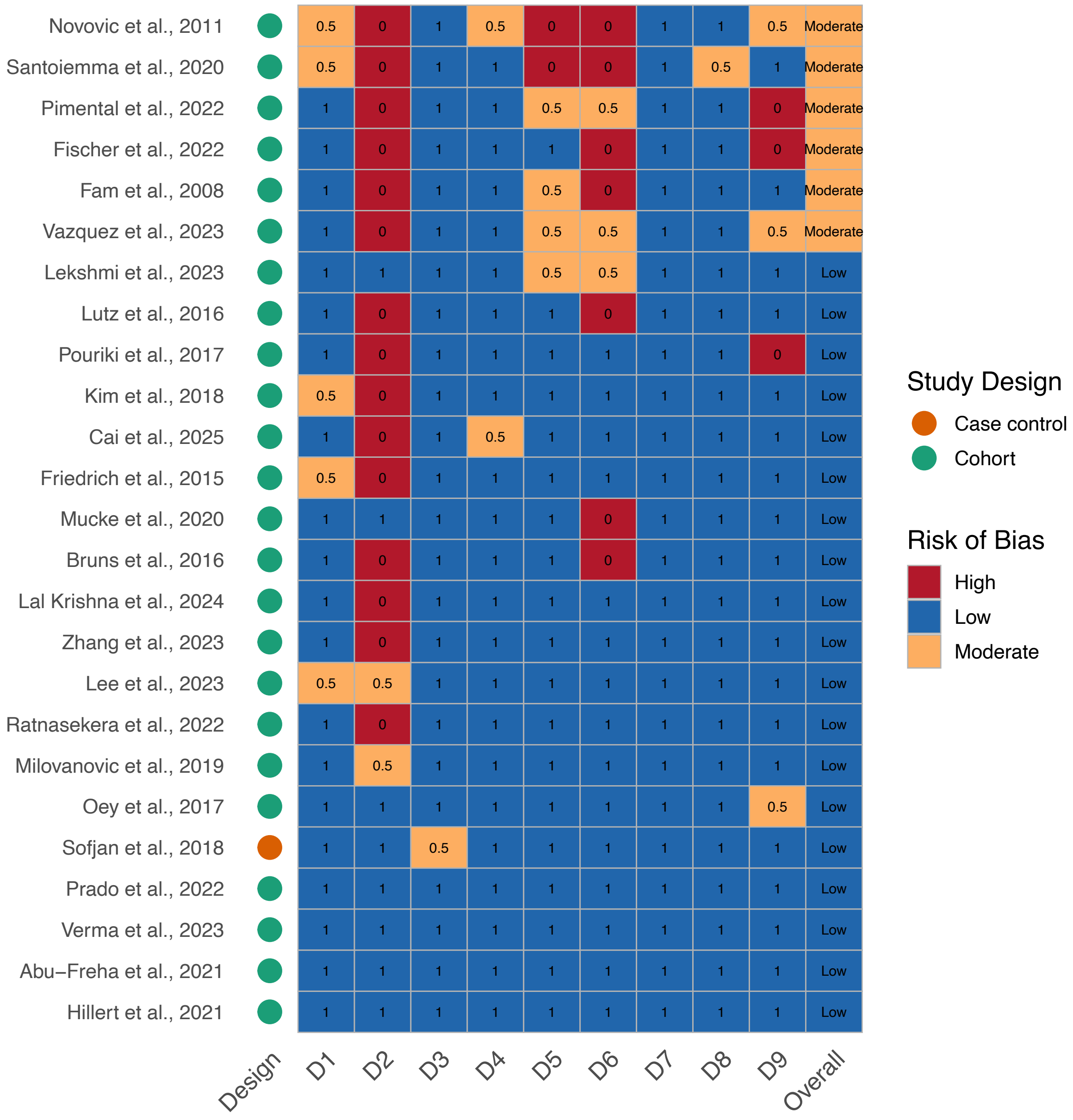

**Figure 1. Traffic-style visualization of study design and risk of bias across cohort and case-control studies.** Each row represents a study, with the first column showing study design (circles: green = cohort, orange = case-control) and subsequent columns showing risk of bias domains (D1–D9) and the overall score (tiles colored by risk category: blue = low, yellow = moderate, red = high). Numerical scores within the tiles indicate the assigned value per domain. This layout allows rapid assessment of study design and methodological quality across included studies.

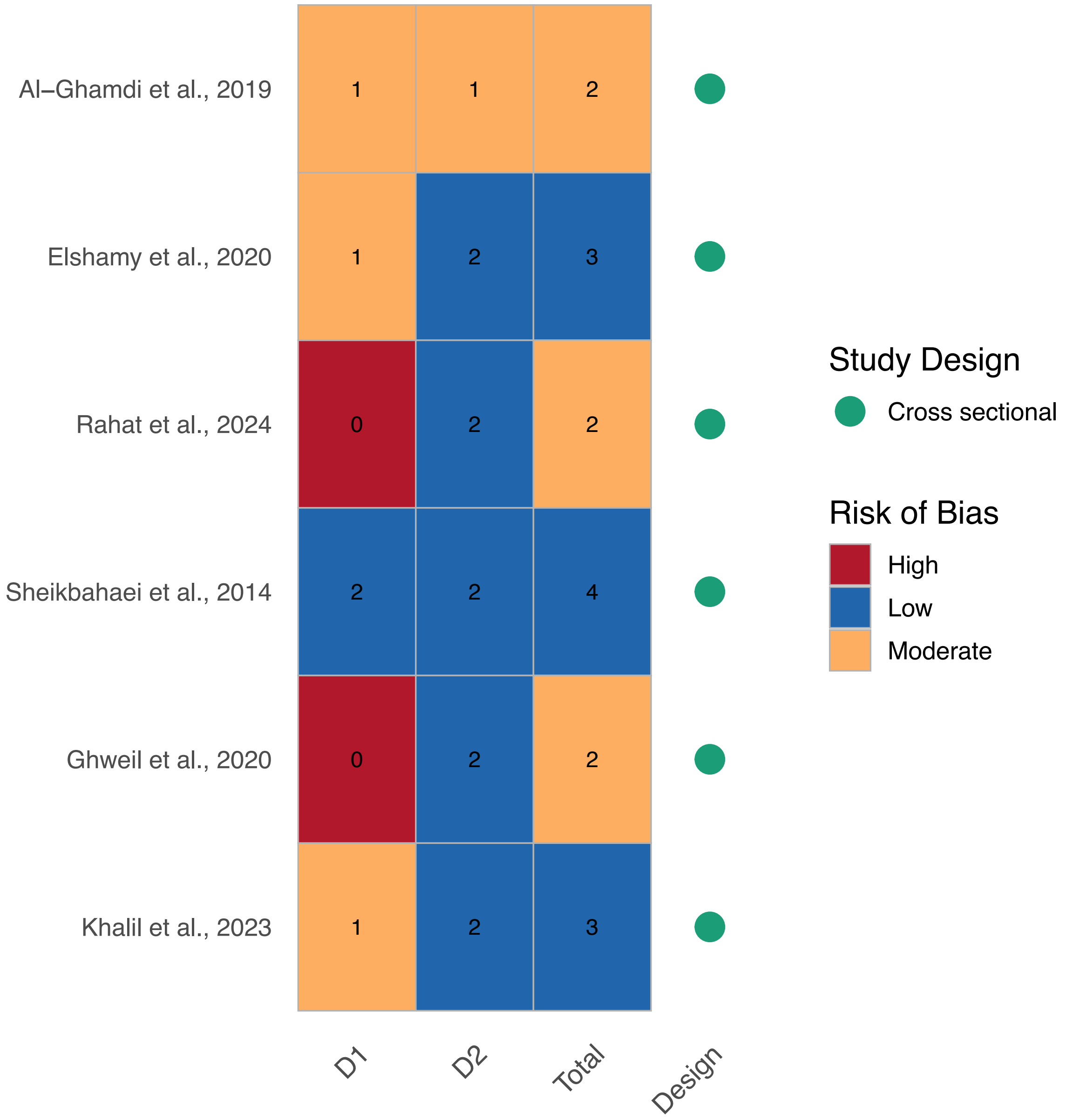

**Figure 2. Traffic-style visualization of study design and risk of bias in cross-sectional studies.** Each row represents a study, with the first three columns showing risk of bias domains D1 and D2, as well as the total score (tiles colored by risk category: blue = low, yellow = moderate, red = high), and the last column indicating study design (green circle = cross-sectional). Numerical values within the tiles indicate the assigned score for each domain or the total.

| Study | Events | Total | GLMM, Random, 95% CI |
|---|---|---|---|
| Abu−Freha et al., 2021 | 31 | 77 | 0.40 [0.29; 0.52] |
| Hillert et al., 2021 | 29 | 160 | 0.18 [0.12; 0.25] |
| Ratnasekera et al., 2022 | 142 | 1019 | 0.14 [0.12; 0.16] |
| Kim et al., 2018 | 77 | 188 | 0.41 [0.34; 0.48] |
| Santoiemma et al., 2020 | 28 | 112 | 0.25 [0.17; 0.34] |
| Al−Ghamdi et al., 2019 | 41 | 103 | 0.40 [0.30; 0.50] |
| Fam et al., 2008 | 34 | 71 | 0.48 [0.36; 0.60] |
| Pimental et al., 2022 | 22 | 65 | 0.34 [0.23; 0.47] |
| Zhang et al., 2023 | 96 | 1171 | 0.08 [0.07; 0.10] |
| Milovanovic et al., 2019 | 6 | 65 | 0.09 [0.03; 0.19] |
| Elshamy et al., 2020 | 21 | 35 | 0.60 [0.42; 0.76] |
| Rahat et al., 2024 | 22 | 35 | 0.63 [0.45; 0.79] |
| Vazquez et al., 2023 | 68 | 271 | 0.25 [0.20; 0.31] |
| Sofjan et al., 2018 | 18 | 47 | 0.38 [0.25; 0.54] |
| Oey et al., 2017 | 22 | 58 | 0.38 [0.26; 0.52] |
| Sheikbahaei et al., 2014 | 106 | 291 | 0.36 [0.31; 0.42] |
| Prado et al., 2022 | 78 | 319 | 0.24 [0.20; 0.30] |
| Mucke et al., 2020 | 18 | 79 | 0.23 [0.14; 0.34] |
| Lutz et al., 2016 | 11 | 49 | 0.22 [0.12; 0.37] |
| Friedrich et al., 2015 | 35 | 128 | 0.27 [0.20; 0.36] |
| Novovic et al., 2011 | 31 | 187 | 0.17 [0.12; 0.23] |
| Lal Krishna et al., 2024 | 16 | 44 | 0.36 [0.22; 0.52] |
| Arya Lekshmi et al., 2023 | 20 | 31 | 0.65 [0.45; 0.81] |
| Lee et al., 2023 | 19 | 59 | 0.32 [0.21; 0.46] |
| Bruns et al., 2016 | 4 | 25 | 0.16 [0.05; 0.36] |
| Fischer et al., 2022 | 20 | 124 | 0.16 [0.10; 0.24] |
| Ghweil et al., 2020 | 17 | 51 | 0.33 [0.21; 0.48] |
| Khalil et al., 2023 | 36 | 84 | 0.43 [0.32; 0.54] |
| Pouriki et al., 2017 | 18 | 60 | 0.30 [0.19; 0.43] |
| **Total (95% CI)** | | **5008** | **0.29 [0.24; 0.35]** |
| **Prediction interval** | | | **[0.09; 0.63]** |

Heterogeneity: $Tau^2 = 0.4607$; $Chi^2 = 417.45$, df = 28 ($P < 0.0001$); $I^2 = 93.3\%$

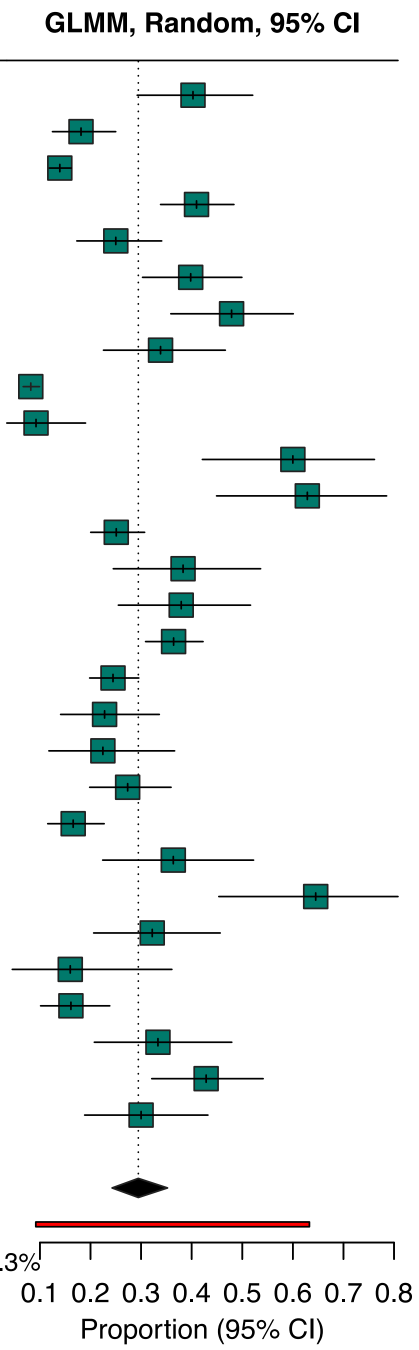

**Supplementary Figure 3.** Pooled proportion of *Escherichia coli* isolates among cirrhosis-associated non-bacteraemic infections. Forest plots display study-level prevalence (squares) with 95% confidence intervals, stratified by Gram status. Diamonds indicate pooled random-effects estimates with 95% confidence and prediction intervals.

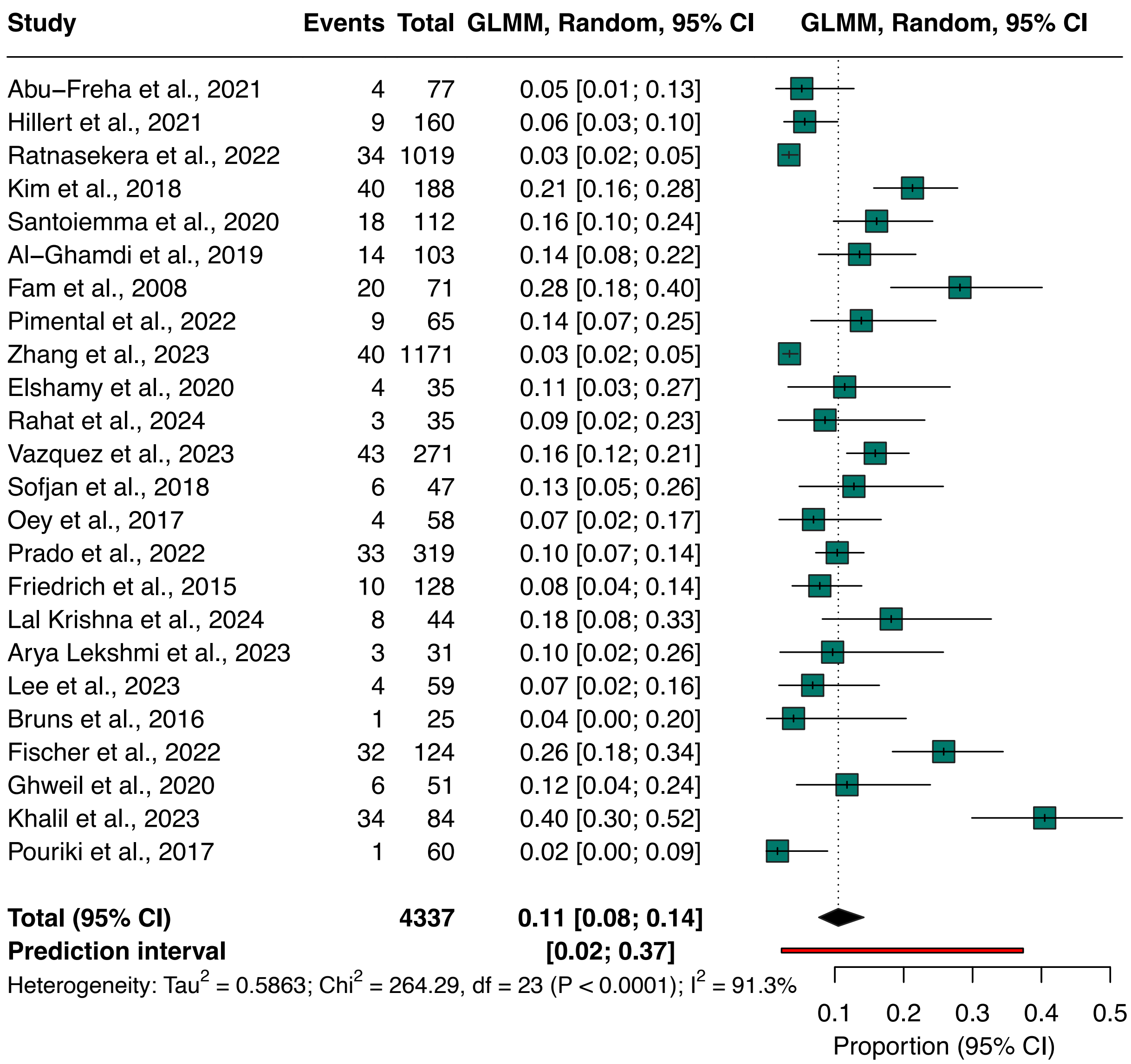

**Supplementary Figure 4.** Pooled proportion of *Klebsiella pneumoniae* isolates among cirrhosis-associated non-bacteraemic infections. Forest plots display study-level prevalence (squares) with 95% confidence intervals, stratified by Gram status. Diamonds indicate pooled random-effects estimates with 95% confidence and prediction intervals.

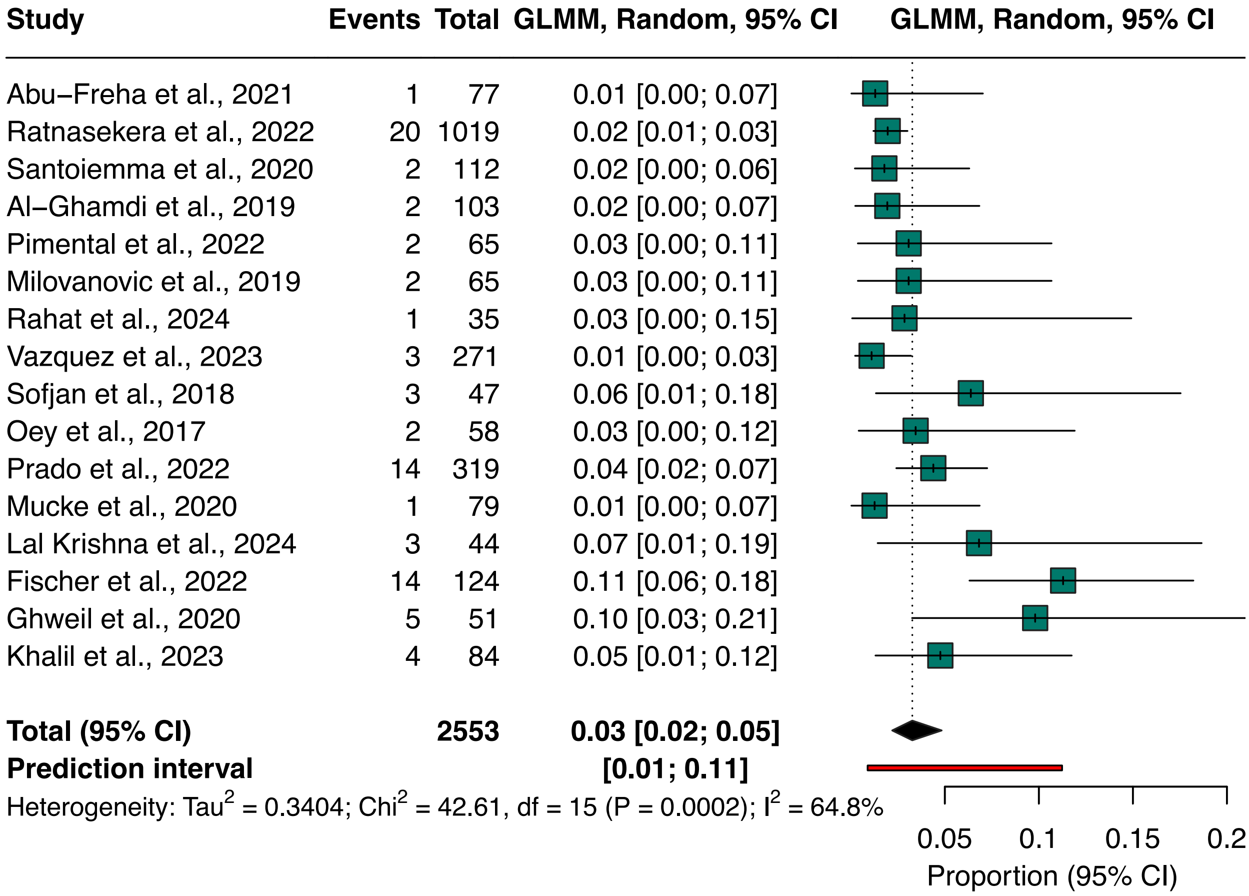

**Supplementary Figure 5.** Pooled proportion of *Pseudomonas aeruginosa* isolates among cirrhosis-associated non-bacteraemic infections. Forest plots display study-level prevalence (squares) with 95% confidence intervals, stratified by Gram status. Diamonds indicate pooled random-effects estimates with 95% confidence and prediction intervals.

| Study | Events | Total | GLMM, Random, 95% CI |
|---|---|---|---|
| Abu−Freha et al., 2021 | 7 | 77 | 0.09 [0.04; 0.18] |
| Hillert et al., 2021 | 17 | 160 | 0.11 [0.06; 0.16] |
| Ratnasekera et al., 2022 | 79 | 1019 | 0.08 [0.06; 0.10] |
| Kim et al., 2018 | 5 | 188 | 0.03 [0.01; 0.06] |
| Santoiemma et al., 2020 | 3 | 112 | 0.03 [0.01; 0.08] |
| Al−Ghamdi et al., 2019 | 4 | 103 | 0.04 [0.01; 0.10] |
| Fam et al., 2008 | 2 | 71 | 0.03 [0.00; 0.10] |
| Pimental et al., 2022 | 5 | 65 | 0.08 [0.03; 0.17] |
| Zhang et al., 2023 | 10 | 1171 | 0.01 [0.00; 0.02] |
| Milovanovic et al., 2019 | 3 | 65 | 0.05 [0.01; 0.13] |
| Elshamy et al., 2020 | 4 | 35 | 0.11 [0.03; 0.27] |
| Rahat et al., 2024 | 4 | 35 | 0.11 [0.03; 0.27] |
| Vazquez et al., 2023 | 28 | 271 | 0.10 [0.07; 0.15] |
| Sofjan et al., 2018 | 4 | 47 | 0.09 [0.02; 0.20] |
| Oey et al., 2017 | 4 | 58 | 0.07 [0.02; 0.17] |
| Sheikbahaei et al., 2014 | 28 | 291 | 0.10 [0.06; 0.14] |
| Mucke et al., 2020 | 8 | 79 | 0.10 [0.04; 0.19] |
| Friedrich et al., 2015 | 5 | 128 | 0.04 [0.01; 0.09] |
| Novovic et al., 2011 | 15 | 187 | 0.08 [0.05; 0.13] |
| Arya Lekshmi et al., 2023 | 1 | 31 | 0.03 [0.00; 0.17] |
| Bruns et al., 2016 | 1 | 25 | 0.04 [0.00; 0.20] |
| Fischer et al., 2022 | 2 | 124 | 0.02 [0.00; 0.06] |
| Ghweil et al., 2020 | 11 | 51 | 0.22 [0.11; 0.35] |
| Khalil et al., 2023 | 4 | 84 | 0.05 [0.01; 0.12] |
| **Total (95% CI)** | | **4477** | **0.06 [0.04; 0.08]** |
| **Prediction interval** | | | **[0.01; 0.21]** |

Heterogeneity: $Tau^2 = 0.4702$; $Chi^2 = 95.97$, df = 23 ($P < 0.0001$); $I^2 = 76.0\%$

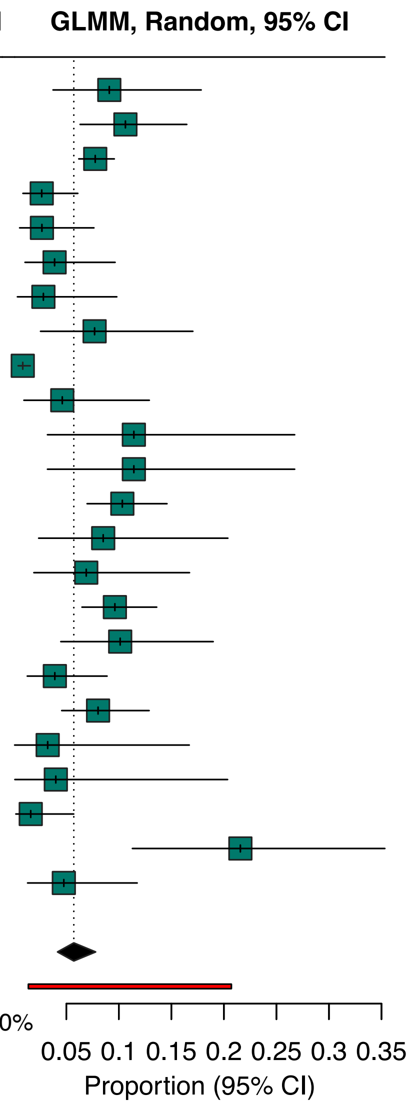

**Supplementary Figure 6.** Pooled proportion of *Staphylococcus aureus* isolates among cirrhosis-associated non-bacteraemic infections. Forest plots display study-level prevalence (squares) with 95% confidence intervals, stratified by Gram status. Diamonds indicate pooled random-effects estimates with 95% confidence and prediction intervals.

| Study | Events | Total | GLMM, Random, 95% CI |
|---|---|---|---|
| Abu−Freha et al., 2021 | 6 | 77 | 0.08 [0.03; 0.16] |
| Hillert et al., 2021 | 40 | 160 | 0.25 [0.19; 0.32] |
| Ratnasekera et al., 2022 | 74 | 1019 | 0.07 [0.06; 0.09] |
| Santoiemma et al., 2020 | 15 | 112 | 0.13 [0.08; 0.21] |
| Al−Ghamdi et al., 2019 | 3 | 103 | 0.03 [0.01; 0.08] |
| Fam et al., 2008 | 5 | 71 | 0.07 [0.02; 0.16] |
| Pimental et al., 2022 | 7 | 65 | 0.11 [0.04; 0.21] |
| Zhang et al., 2023 | 42 | 1171 | 0.04 [0.03; 0.05] |
| Milovanovic et al., 2019 | 34 | 65 | 0.52 [0.40; 0.65] |
| Elshamy et al., 2020 | 2 | 35 | 0.06 [0.01; 0.19] |
| Vazquez et al., 2023 | 21 | 271 | 0.08 [0.05; 0.12] |
| Sofjan et al., 2018 | 4 | 47 | 0.09 [0.02; 0.20] |
| Oey et al., 2017 | 5 | 58 | 0.09 [0.03; 0.19] |
| Sheikbahaei et al., 2014 | 27 | 291 | 0.09 [0.06; 0.13] |
| Prado et al., 2022 | 158 | 319 | 0.50 [0.44; 0.55] |
| Mucke et al., 2020 | 26 | 79 | 0.33 [0.23; 0.44] |
| Lutz et al., 2016 | 7 | 49 | 0.14 [0.06; 0.27] |
| Friedrich et al., 2015 | 36 | 128 | 0.28 [0.21; 0.37] |
| Novovic et al., 2011 | 22 | 187 | 0.12 [0.08; 0.17] |
| Lal Krishna et al., 2024 | 4 | 44 | 0.09 [0.03; 0.22] |
| Lee et al., 2023 | 10 | 59 | 0.17 [0.08; 0.29] |
| Bruns et al., 2016 | 12 | 25 | 0.48 [0.28; 0.69] |
| Fischer et al., 2022 | 27 | 124 | 0.22 [0.15; 0.30] |
| Ghweil et al., 2020 | 2 | 51 | 0.04 [0.00; 0.13] |
| Pouriki et al., 2017 | 24 | 60 | 0.40 [0.28; 0.53] |
| **Total (95% CI)** | | **4670** | **0.14 [0.10; 0.20]** |
| **Prediction interval** | | | **[0.02; 0.56]** |

Heterogeneity: $Tau^2 = 0.9606$; $Chi^2 = 557.74$, df = 24 ($P < 0.0001$); $I^2 = 95.7\%$

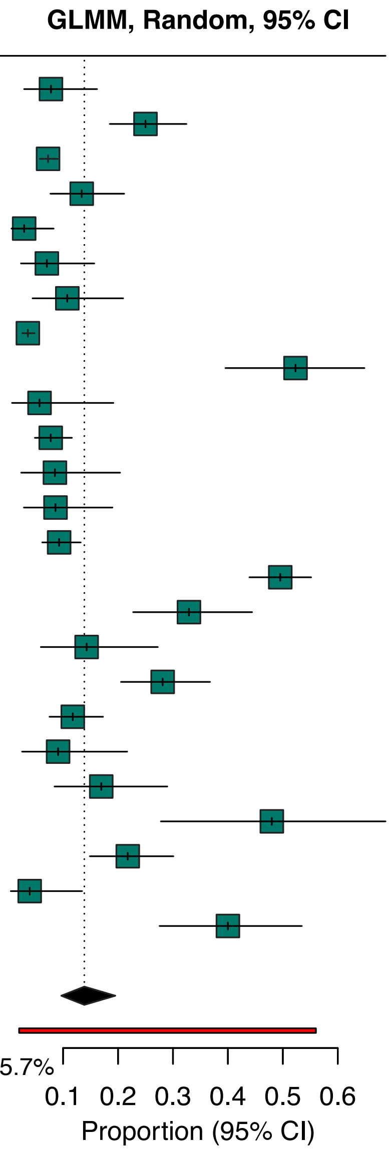

**Supplementary Figure 7.** Pooled proportion of Enterococci isolates among cirrhosis-associated non-bacteraemic infections. Forest plots display study-level prevalence (squares) with 95% confidence intervals, stratified by Gram status. Diamonds indicate pooled random-effects estimates with 95% confidence and prediction intervals.

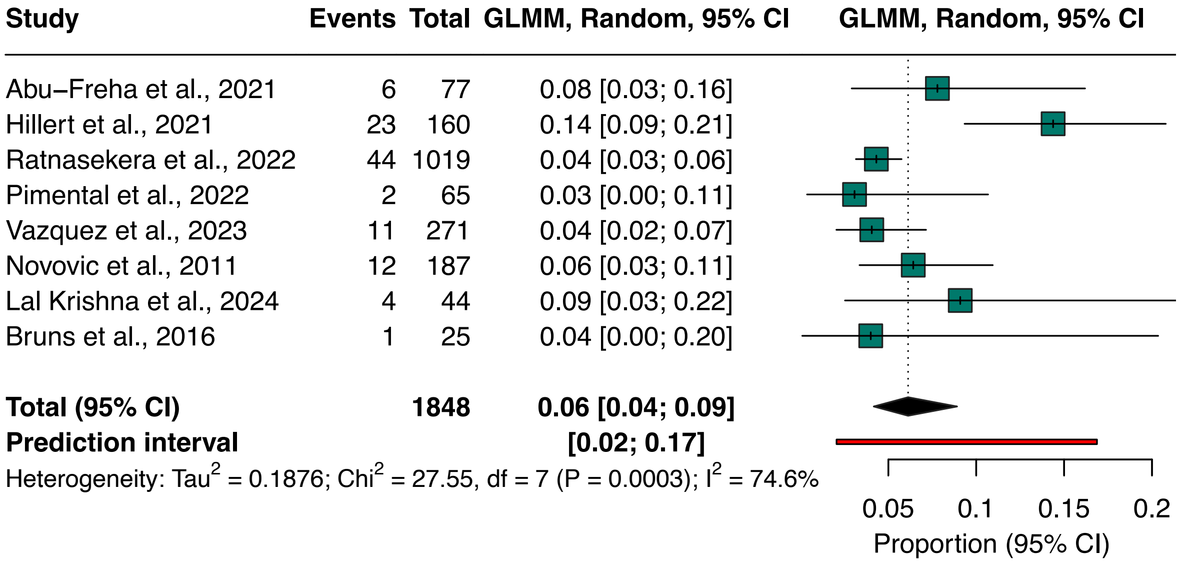

**Supplementary Figure 8.** Pooled proportion of *Enterococcus faecalis* isolates among cirrhosis-associated non-bacteraemic infections. Forest plots display study-level prevalence (squares) with 95% confidence intervals, stratified by Gram status. Diamonds indicate pooled random-effects estimates with 95% confidence and prediction intervals.

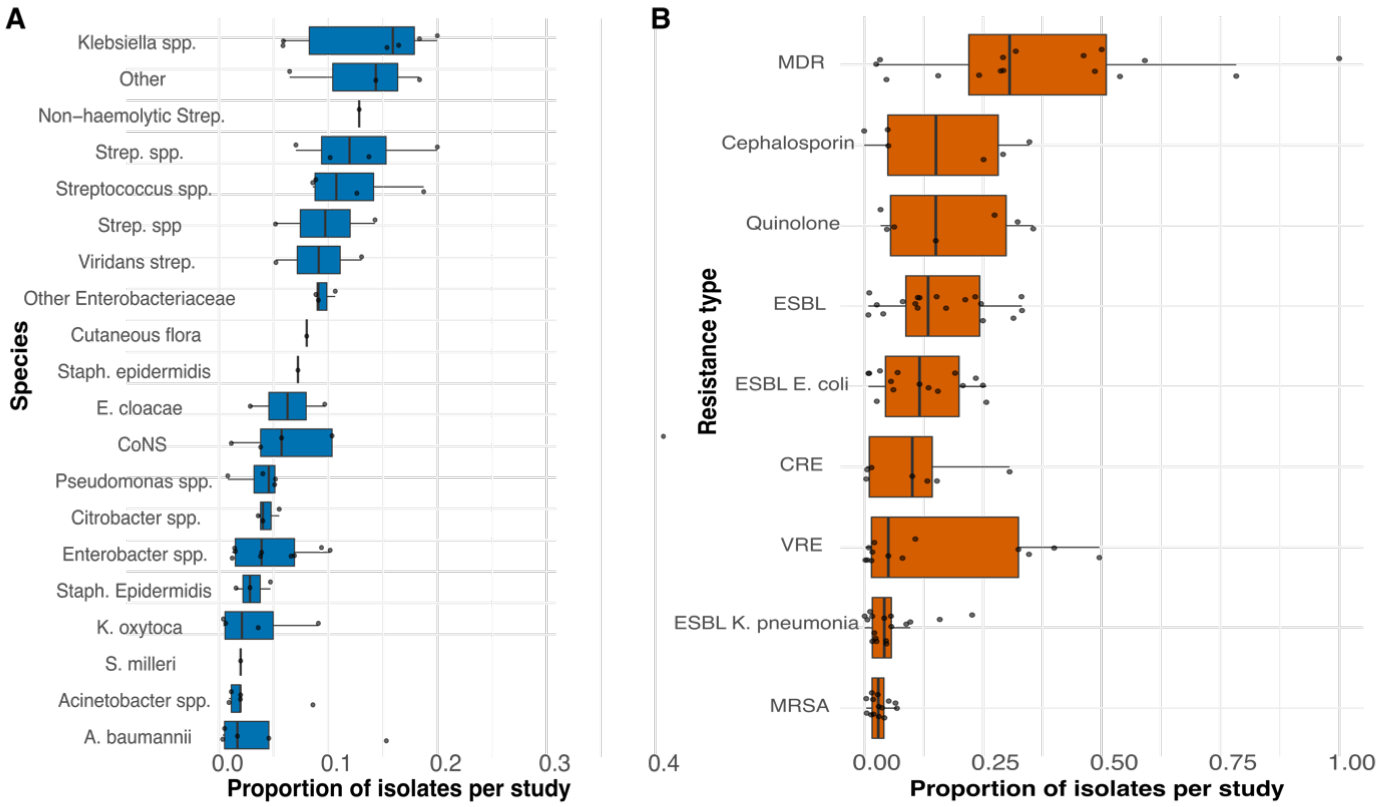

**Supplementary Figure 9: Study-level distributions of bacterial species and antimicrobial resistance phenotypes in cirrhosis-associated non-bacteraemic infections. (A)** Distribution of the top 20 bacterial species across studies. Boxplots show study-level proportions of the 20 most frequently reported bacterial species, with individual points representing study-specific estimates, illustrating heterogeneity in species prevalence. Species are ordered by median proportion. **(B)** Distribution of major antimicrobial resistance phenotypes across studies. Boxplots show study-level proportions for MRSA, ESBL (overall and species-specific), VRE, CRE, fluoroquinolone resistance, extended-spectrum cephalosporin resistance, and multidrug resistance, with individual points representing study-specific estimates.
